# Uncovering the multivariate genetic architecture of frailty with genomic structural equation modelling

**DOI:** 10.1101/2024.07.24.24310923

**Authors:** Isabelle F Foote, Jonny P Flint, Anna E Fürtjes, Donncha S Mullin, John D Fisk, Tobias K Karakach, Andrew Rutenberg, Nicholas G Martin, Michelle K Lupton, David J Llewellyn, Janice M Ranson, Simon R Cox, Michelle Luciano, Kenneth Rockwood, Andrew D Grotzinger

## Abstract

Frailty is a multifaceted clinical state associated with accelerated aging and adverse health outcomes. Informed etiological models of frailty hold promise for producing widespread health improvements across the aging population. Frailty is currently measured using aggregate scores, which obscure etiological pathways that are only relevant to subcomponents of frailty. Therefore, we performed the first multivariate genome-wide association study of the latent genetic architecture between 30 frailty deficits, which identified 408 genomic risk loci. Our model included a general factor of genetic overlap across all deficits, plus six novel factors indexing shared genetic signal across specific groups of deficits. Follow-up analyses demonstrated the added clinical and etiological value of the six factors, including predicting frailty in external datasets, divergent genetic correlations with clinically relevant outcomes, and unique underlying biology linked to aging. This suggests nuanced models of frailty are key to understanding its causes and how it relates to worse health.

---

Frailty is a complex clinical state that affects more than 40% of adults aged over 65 years(1). It is defined as a state of progressive, multisystem physiological decline that reduces an individual’s ability to withstand external stressors(1). This deterioration can lead to both physical and mental impairment and is strongly associated with adverse health outcomes, including earlier mortality(2) and increased levels of disability and hospitalization(3). Global population aging means that frailty represents a growing public health concern(4). Family-based studies indicate a substantial genetic component to frailty, with heritability estimates of ∼45%(5). Therefore, genetic methods offer a promising tool for better understanding the risk pathways to this critical health state. Nevertheless, the etiology of frailty remains largely unknown, limiting our potential to identify effective therapeutic or preventive treatments. We hypothesized that what has limited our etiological understanding is that we have traditionally only considered aggregate measurements of frailty in genetic studies.

The two most common methods for measuring frailty are the Frailty Index (FI) and Fried Frailty Score (FFS)(6, 7). The FI quantifies frailty by calculating the proportion of ‘deficits’ that are present within an individual from a set of ≥30 phenotypes associated with poor health outcomes in older adults(6). The FFS uses an aggregate score across five physical frailty deficits (weight loss, weakness, exhaustion, slow walking speed and physical inactivity), where the presence of ≥3 of these deficits indicates frailty(7). Although the FFS is easier to measure in large samples, it is intended to capture pathways of physical frailty and does not provide sufficient information to assess the more nuanced subgroupings that may occur within the broader frailty construct(8). By contrast, the deficits that form the FI span many levels of functional, psychological, and social aspects of health, allowing frailty to be measured across a broad spectrum of traits. Yet, FI deficits are heterogeneous and vary in their underlying etiology.

Phenotypic work clearly indicates that the deficits comprising the FI are not always strongly correlated and are driven by diverse biological mechanisms(9–11). Their combination into a single aggregate score is, therefore, likely to obscure causal pathways of frailty. For example, previous work that applied principal component analysis to phenotypic data of FI deficits demonstrated that additional informative variance associated with frailty was captured when three clusters were modelled instead of one cluster(12). Information that is lost by aggregation could be identified by a more detailed genetic analysis. Recent advances in multivariate genomics, such as the development of Genomic Structural Equation Modelling (Genomic SEM)(13), offer the opportunity to model the genetic basis of frailty at a multi-dimensional level.

We used Genomic SEM to identify novel groupings of genetic overlap between 30 frailty deficits using publicly available GWAS summary statistics. We identified seven distinct latent factors underpinning frailty that displayed unique genetic overlap with clinically relevant health outcomes and were defined by divergent sets of genomic risk loci and biological pathways, which provided empirical support for a majority of the 12 ‘hallmarks of aging’(14). In combination, these latent constructs yielded enhanced prediction of frailty status in external cohorts and a dramatic improvement in genomic locus discovery compared to aggregate measures.

## Results

### Multivariate Genetic Architecture of Frailty

Following careful quality control and selection of frailty deficit phenotypes (**Online Methods**), we used Genomic SEM to model the genome-wide genetic overlap from GWAS data for 30 frailty deficits(15–19) (**Figure 1** and **Tables S1-S2**). As Genomic SEM can model genetic overlap across even mutually exclusive participant samples, this allowed us to bring together the most well-powered genomic studies currently available for the 30 frailty deficits and produce estimates with the highest possible precision. Using a combination of exploratory and confirmatory factor analysis (**Online Methods**), this modeling procedure yielded a bifactor model that provided a good fit to the data (**Figure 1** and **Table S3**; comparative fit index (CFI) = 0.93; standardized root mean squared residual (SRMR) = 0.07). This model included a General Factor that indexed genetic overlap across all 30 deficits and so is conceptually similar to prior aggregate measures of frailty. In addition, the model produced six residual group factors that were orthogonal (i.e. uncorrelated) to the General Factor, which were defined by additional genetic overlap within subsets of frailty deficits. These six factors captured distinct, albeit inter-correlated, frailty pathways related to social isolation (Factor 1), unhealthy lifestyle (Factor 2), multimorbidity (Factor 3), respiratory/metabolic problems (Factor 4), poorer cognition (Factor 5), and disability (Factor 6). The remaining analyses evaluated the divergent validity and clinical utility of these six factors by examining their ability to uniquely capture frailty-relevant pathways at increasingly granular levels of biological analysis.

**Figure 1:**
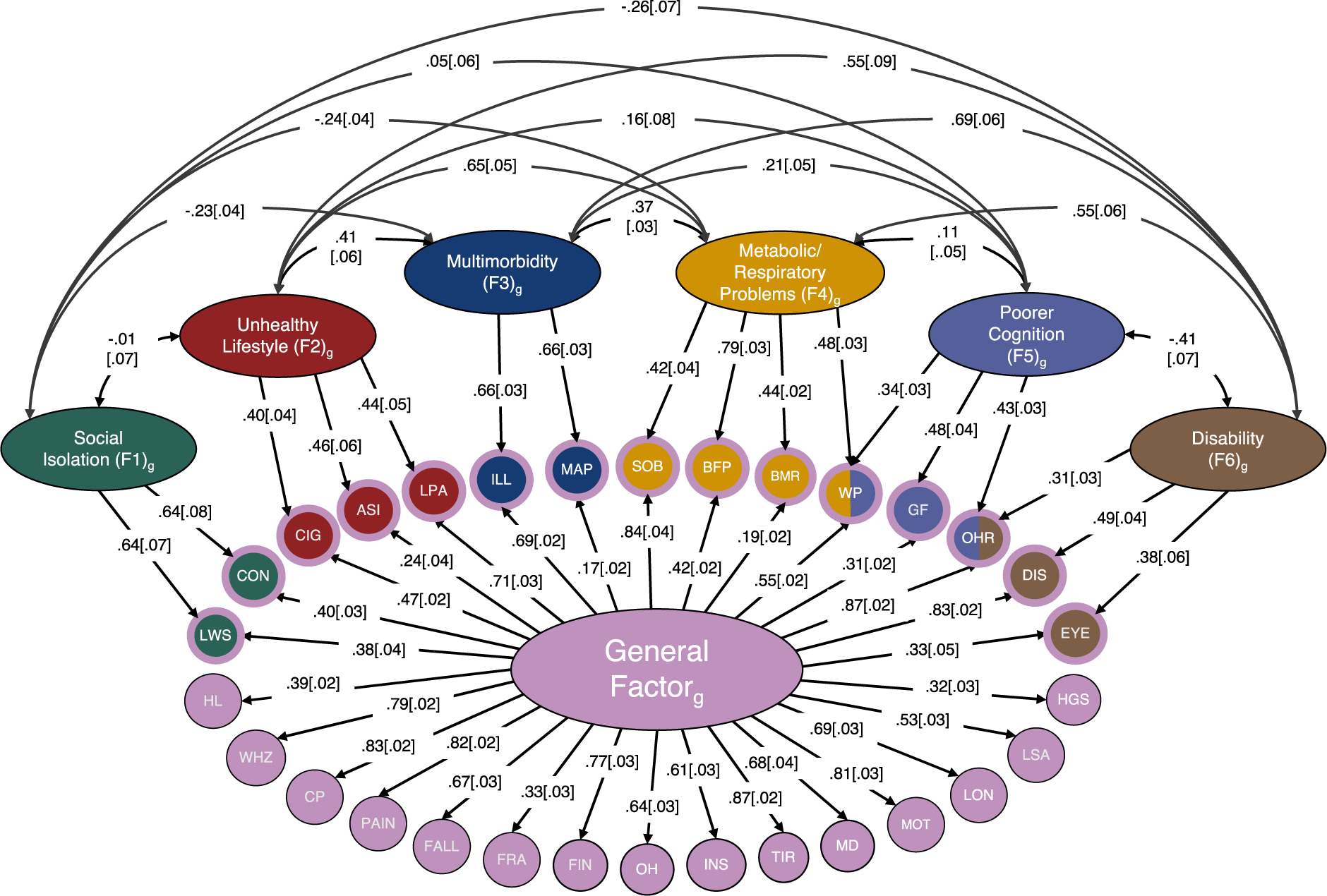
A path diagram of the standardized results for our bifactor model of frailty. All 30 frailty deficits load onto the General Factor of frailty (large pink oval), which is orthogonal to Factors 1 to 6 (i.e. residual factors). The small circles represent the 30 measured frailty indicators (i.e. genetic variance captured in the univariate GWAS for that phenotype), whereas the medium-size ovals represent latent factors (i.e. unmeasured constructs representing genetic overlap between the indicators that load onto them). Single-headed arrows represent a directional genetic correlation between a latent factor and an indicator (i.e. factor loadings), whereas curved double-headed arrows represent inter-factor correlations between Factors 1 to 6. Standard errors of the correlation coefficients are reported in square brackets. SOB = shortness of breath walking on flat ground; BFP = body fat percentage; BMR = basal metabolic rate; WP = slow walking pace; GF = low fluid intelligence score; OHR = poorer overall health rating; DIS = long-standing illness, disability or infirmity; EYE = eye disorder/problem; HGS = low hand grip strength; LSA = low social/leisure activity; LON = loneliness, isolation; MOT = feelings of unenthusiasm/disinterest; MD = major depressive disorder; TIR = tiredness/lethargy; INS = insomnia; OH = poor oral health; FIN = financial difficulties; FRA = fracture in last 5 years; FALL = number of falls in past year; PAIN = pain experienced in past month; CP = chest pain; WHZ = wheeze/whistling in chest in past year; HL = age-related hearing loss; LWS = not living with spouse/partner; CON = unable to confide; CIG = number of cigarettes smoked per day; ASI = pulse wave arterial stiffness index; LPA = physical inactivity; ILL = number of non-cancer illnesses; MAP = mean arterial pressure.

### Genetic Overlap between Latent Frailty Factors and Aging-Related Health Traits

To validate whether the latent factors in the bifactor model reflect different aspects of frailty, we measured the level of genetic correlation between each of the latent factors and 52 aging-related health outcomes and established frailty measures (**Figure 2** and **Tables S4-S5**). The General Factor displayed high positive genetic correlations with the FI (*r*_*g*_ = 0.93 [SE = 0.02], *P*_FDR_ = 9.1×10^−298^) and FFS (*r*_*g*_ = 0.83 [0.02], *P*_FDR_ = 9.1×10^−298^), indicating that the General Factor closely approximates both of these frailty phenotypes in line with expectations, given that this latent factor represents genetic overlap between all 30 included deficits. These findings were corroborated by the fact that all the latent factors (except Factor 1 (social isolation)) were associated with shorter parental lifespan (*r*_*g*_ < −0.31, *P*_FDR_ < 3.2×10^−06^), reduced longevity (*r*_*g*_ < −0.21, *P*_FDR_ < 0.01), increased risk of common aging-related infections (*r*_*g*_ > 0.22, *P*_FDR_ < 0.01), hospitalization due to infection (*r*_*g*_ > 0.15, *P*_FDR_ < 0.008), and heart failure (*r*_*g*_ > 0.15, *P*_FDR_ < 0.005), all of which reflect key clinical correlates of frailty.

**Figure 2:**
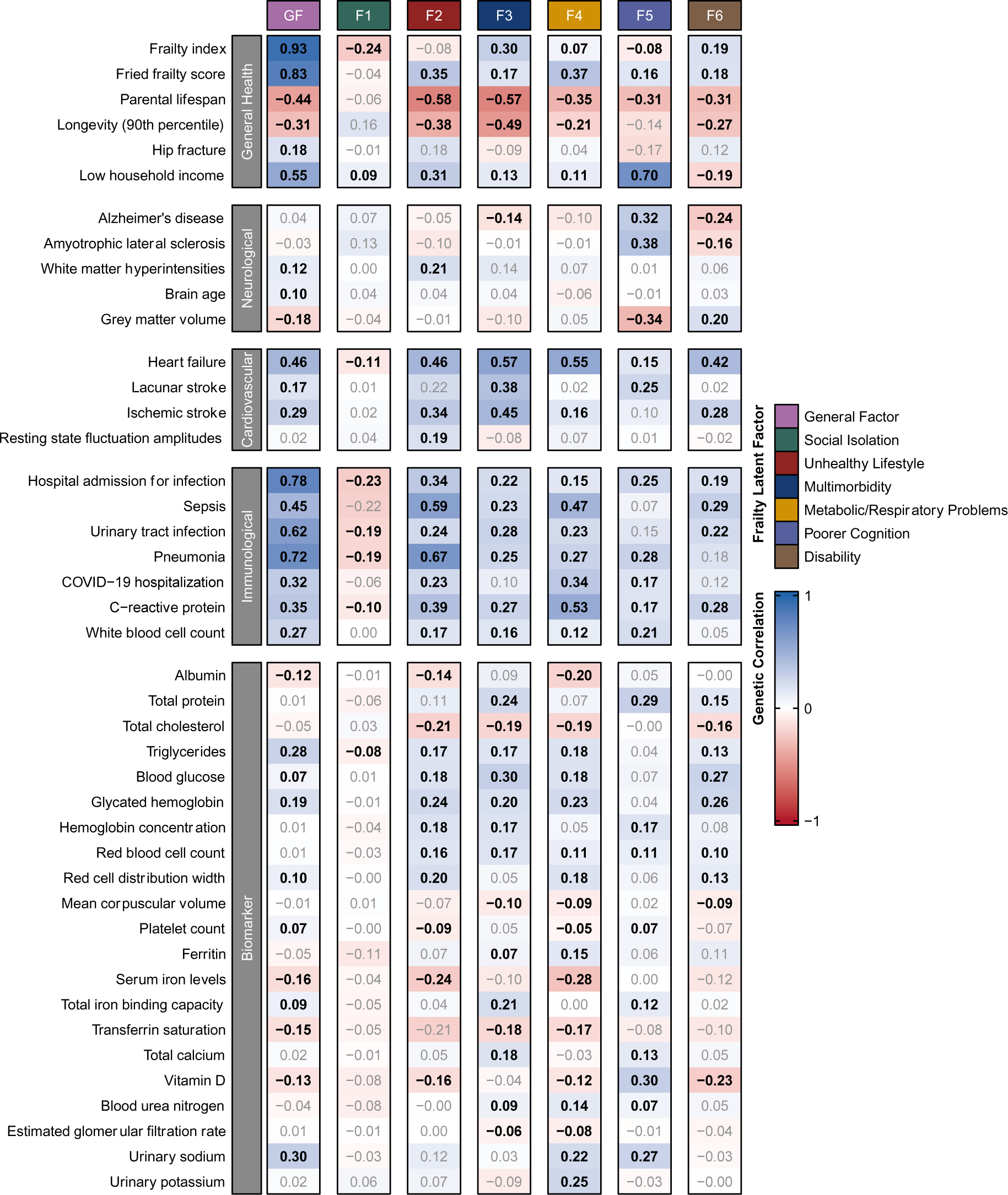
A heatmap of the genetic correlations between aging-related health outcomes and each of the latent factors from the frailty bifactor model. Genetic correlations with an FDR-corrected *P*-value <0.05 are highlighted in bold. Blue shading represents a positive genetic correlation whereas red shading represents a negative genetic correlation. For visualization purposes, only health outcomes that demonstrated at least one FDR-corrected *P*-value <0.05 with one or more of the latent factors are included in this figure (full results can be found in **Table S5**).

Importantly, our findings support the inclusion of additional subgroups when measuring frailty accumulation by highlighting divergent genetic correlations with aging-related health outcomes. For example, Factor 5 (poorer cognition), was the only latent factor to demonstrate positive genetic correlations with Alzheimer’s disease (*r*_*g*_ = 0.32 [SE = 0.07], *P*_FDR_ = 2.26×10^−05^) and amyotrophic lateral sclerosis (*r*_*g*_ = 0.38 [0.06], *P*_FDR_ = 1.07×10^−08^). Factor 5 also captured genetic variance associated with smaller grey matter volume (*r*_*g*_ = −0.34 [0.05], *P*_FDR_ = 9.53×10^−12^) and lacunar stroke (*r*_*g*_ = 0.25 [0.09], *P*_FDR_ = 9.80×10^−03^). By contrast, Factor 2 (unhealthy lifestyle) demonstrated genetic correlations with brain-related vascular changes, including increased white matter hyperintensities (*r*_*g*_ = 0.21 [0.09], *P*_FDR_ = 0.04) and resting state fluctuation amplitudes (*r*_*g*_ = 0.19 [0.08], *P*_FDR_ = 0.03), whereas Factor 3 (multimorbidity) was correlated with ischemic and lacunar stroke (*r*_*g*_ = 0.45 [0.05], *P*_FDR_ = 6.21×10^−18^ and *r*_*g*_ = 0.38 [0.09], *P*_FDR_ = 3.00×10^−03^), but not cerebrovascular markers. Finally, all the latent frailty factors displayed distinct patterns of genetic correlations with routinely collected blood and urinary biomarkers (**Figure 2**), which may represent potential endophenotype profiles for these frailty subgroups. By definition, the identified relationships are independent of shared risk pathways captured by the General Factor of frailty. It follows that our model of frailty has potential clinical utility for targeted prevention and therapeutic intervention in patients that present with elevated risk within a subgroup of frailty deficits.

### Multivariate GWAS Identifies 408 Genomic Risk Loci Associated with Frailty

We subsequently performed a multivariate GWAS of our frailty bifactor model using Genomic SEM to uncover genomic risk loci that were associated with each latent frailty factor (*P_BONF_* < 7.14×10^−09^). We pruned out any significantly heterogenous genetic signal (Q_SNP_) from our GWAS results to ensure that we were only measuring genetic effects that were shared between the deficits that defined that particular latent factor (**Online Methods**). From this shared signal, we identified a total of 408 genomic risk loci across the seven latent frailty factors (**Figure 3** and **Tables S6-12**). We integrated the results from our GWAS with the lead single nucleotide polymorphisms (SNPs) of the risk loci identified in the previously published GWAS of the FI(20) (**Table S13**) and FFS(21) (**Table S14)**. Of the overlapping lead SNPs (12/14 for the FI and 18/37 for the FFS), we replicated 33.3% of the previously identified genomic risk loci for the FI and 22.2% for the FFS with the General Factor. 100% of the risk loci for both frailty phenotypes displayed nominal significance with at least one of our latent frailty factors, supporting validation of our model in measuring frailty. However, 33.3% of the FI loci and 25% of the FFS loci were significant for the SNP-level heterogeneity metric (Q_SNP_ *P* < 7.14×10^−09^; **Tables S13-14**). As Q_SNP_ identifies SNPs that are likely to be deficit-specific or that show heterogeneous effects between the included frailty deficits, these findings indicate that existing aggregate frailty measures are sensitive to large effects driven by a single indicator that are not part of more general frailty pathways. Furthermore, 33.3% and 27.8% of the FI and FFS loci were replicated by the residual factors (i.e. Factors 1 to 6) instead of the General Factor. This demonstrates that our model can comprehensively define subgroups within the frailty construct, which one aggregate measure overlooks.

**Figure 3:**
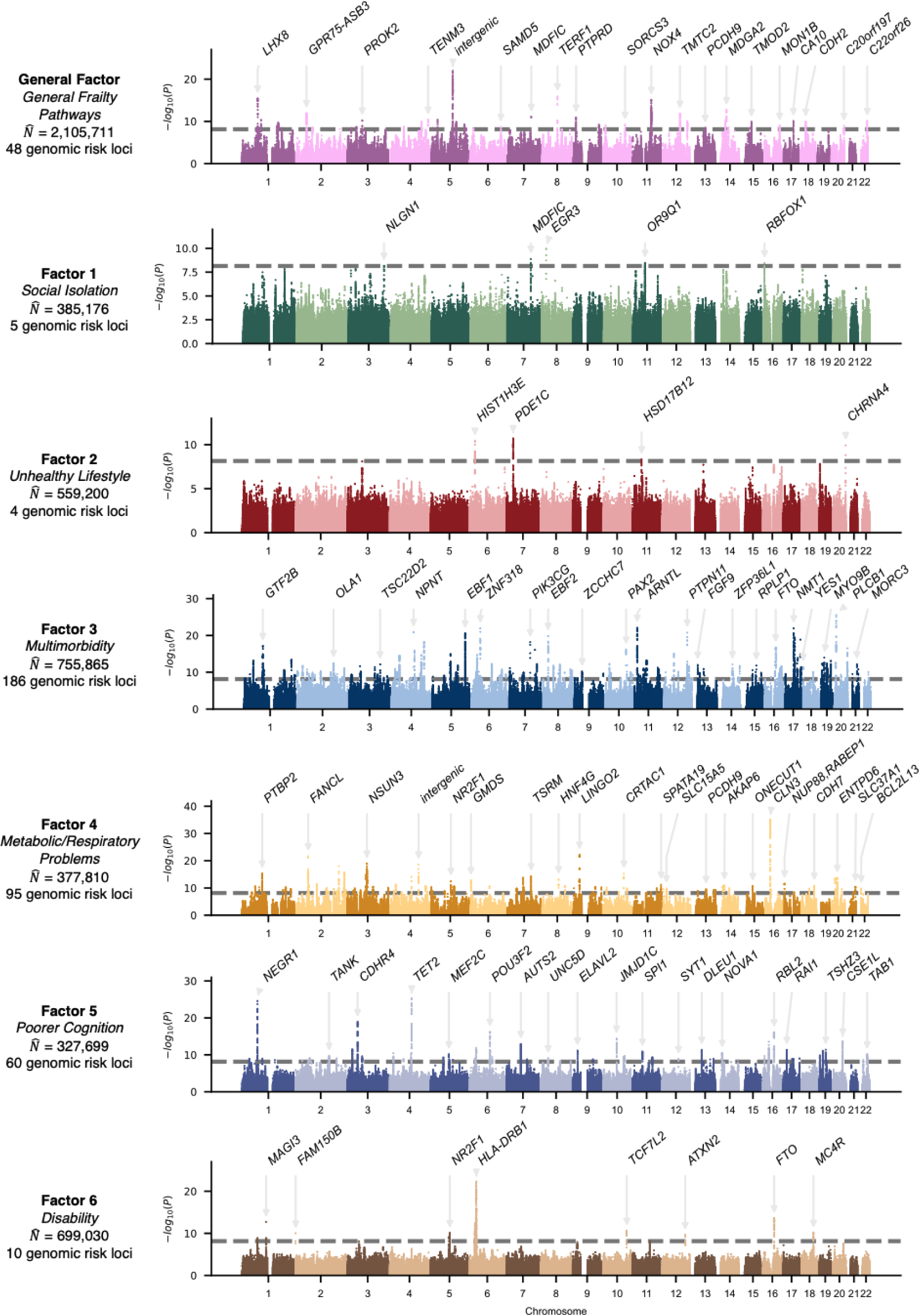
Manhattan plots of the shared genetic signal for each of the latent factors in the frailty model. The x-axis depicts the chromosomes and the y-axis represents the −log^10^ P-values of the association between each individual SNP and each latent factor. The closest gene to the lead SNP are annotated for top loci for each latent factor. The dashed grey line denotes the genome-wide significance threshold adjusted for multiple testing (i.e. P_BONF_ < 7.14×10^−09^). 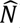 is the expected sample size of each latent factor implied by the GWAS summary statistics of that factor, which are influenced by the power of the factor loadings of the indicators (i.e. frailty deficits) that define it.

### Gene Expression and Epigenetic Changes Associated with Latent Frailty Factors

We completed a series of post-GWAS analyses to explore how the genetics underpinning each of the frailty latent factors may influence underlying biology. We applied Multi-marker Analysis of GenoMic Annotation (MAGMA) gene property analysis to test for enriched gene expression changes in 54 body tissues and Stratified Genomic SEM to test for enrichment in 146 functional annotations linked to gene expression and epigenetic changes in tissues and cell subtypes within the brain (**Figure 4** and **Online Methods**).

**Figure 4:**
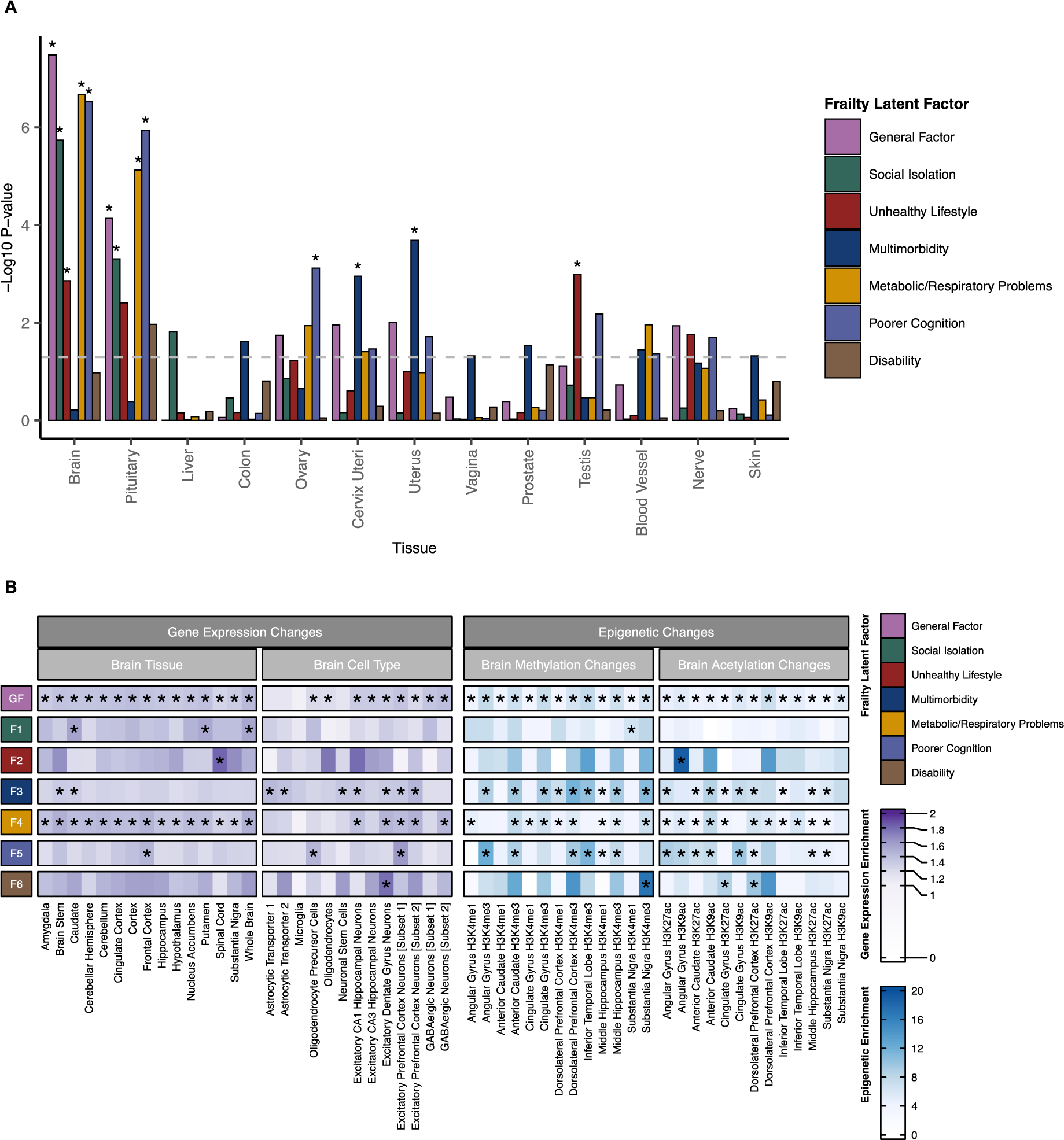
Results from the MAGMA gene property analysis (Panel A) and the Stratified Genomic SEM (Panel B). In *Panel A*, the y-axis denotes the −log^10^ *P*-value of the enrichment between each latent frailty factor and body tissues from GTEx version 8 (only tissues with significant enrichment are displayed). The light grey dashed line denotes the cut-off for nominal significance (i.e. *P-*value < 0.05) and bars that are marked with an asterisk indicate enrichment that remained significant after adjusting for multiple testing (i.e. *P_FDR_* < 0.05). *Panel B* displays heatmaps of the enrichment values calculated using Stratified Genomic SEM to test for differences in gene expression and epigenetic marks associated with each latent frailty factor in a selection of brain-relevant tissues and cell types. Significant enrichment values are marked with an asterisk (*P_FDR_* < 0.05). GF = General Factor; F1 = Factor 1; F2 = Factor 2; F3 = Factor 3; F4 = Factor 4; F5 = Factor 5; F6 = Factor 6).

MAGMA gene property analysis showed significant enrichment in the brain and pituitary gland for all the latent frailty factors except for those underlying multimorbidity (Factor 3) and disability (Factor 6) pathways (**Figure 4A**). The only other tissues that demonstrated significantly enriched changes in gene expression were the reproductive organs, whereby Factor 2 (unhealthy lifestyle) showed significant gene expression changes in the testis, Factor 3 (multimorbidity) showed significant changes in gene expression in the cervix and uterus, and Factor 5 (poorer cognition) showed significant gene expression changes within the ovaries (**Table S15**).

Stratified Genomic SEM provided a more in-depth picture of enrichment in the brain (**Table S16**). We found widespread enrichment in gene expression and epigenetic changes throughout the brain regions in oligodendrocytes and neurons for the General Factor of frailty. However, the widespread nature of this enrichment demonstrates that using an aggregate measure of frailty is less likely to provide a fine-tuned picture of the underlying mechanisms of frailty due to its generalized impact on brain function. In contrast, the residual factors provided a more detailed understanding of pathways implicated by different frailty deficits, which could present future therapeutic targets within the broad spectrum of frailty (**Figure 4B**). For instance, Factor 1 (social isolation) only showed significant gene expression changes in the dorsal striatum (caudate and putamen) and methylation changes in the substantia nigra, whereas Factor 2 (unhealthy lifestyle) showed enriched gene expression in the spinal cord, but not any of the tested brain regions. In addition, Factor 5 (poorer cognition) showed gene expression enrichment in excitatory prefrontal cortex neurons and oligodendrocyte precursor cells, as well as epigenetic changes in the angular gyrus, cingulate gyrus, anterior caudate, dorsolateral prefrontal cortex, hippocampus and substantia nigra.

### Gene Prioritization and Pathway Analysis

We used five methods to map potentially causal genes to each latent frailty factor to assess the biological pathways that might be associated with each frailty subgroup. These methods included mapping SNPs to genes based on their position, whether they were known expression quantitative trait loci (eQTLs) or if they were located in promoter regions known to regulate chromatin interactions (**Tables S17-S23**). In addition, we performed a genome-wide gene-based test using MAGMA (**Tables S24-S30**) and applied summary data-based Mendelian randomization (SMR) to identify SNPs that demonstrated evidence of having a pleiotropic effect on expression, splicing or methylation changes in gene function (**Tables S31-S37**). We triangulated the results from these five gene mapping techniques and prioritized the most likely candidate genes based on whether they were mapped by three or more of the methods. This resulted in 1195 genes being prioritized, which we took forward for pathway analysis (54 for the General Factor; 4 for Factor 1 (social isolation); 20 for Factor 2 (unhealthy lifestyle); 585 for Factor 3 (multimorbidity); 194 for Factor 4 (metabolic/respiratory problems); 266 for Factor 5 (poorer cognition) and 72 for Factor 6 (disability)) (**Supplementary Results**). Using METASCAPE we performed enrichment analysis to identify gene ontology pathways and disease pathways that were significantly associated with the prioritized genes mapped to each latent factor(22). As there can be extensive redundancy between gene sets, we combined highly correlated enriched pathways into clusters, named according to the gene ontology pathway that had the strongest enrichment with the latent frailty factors (**Online Methods** and **Figure 5**). Since the General Factor was orthogonal to the other latent frailty factors, we conducted pathway analysis separately for that factor, but performed a combined analysis for Factors 1 to 6 to account for the potential overlap in implicated gene pathways owing to the presence of inter-factor correlations between these latent residual factors.

**Figure 5:**
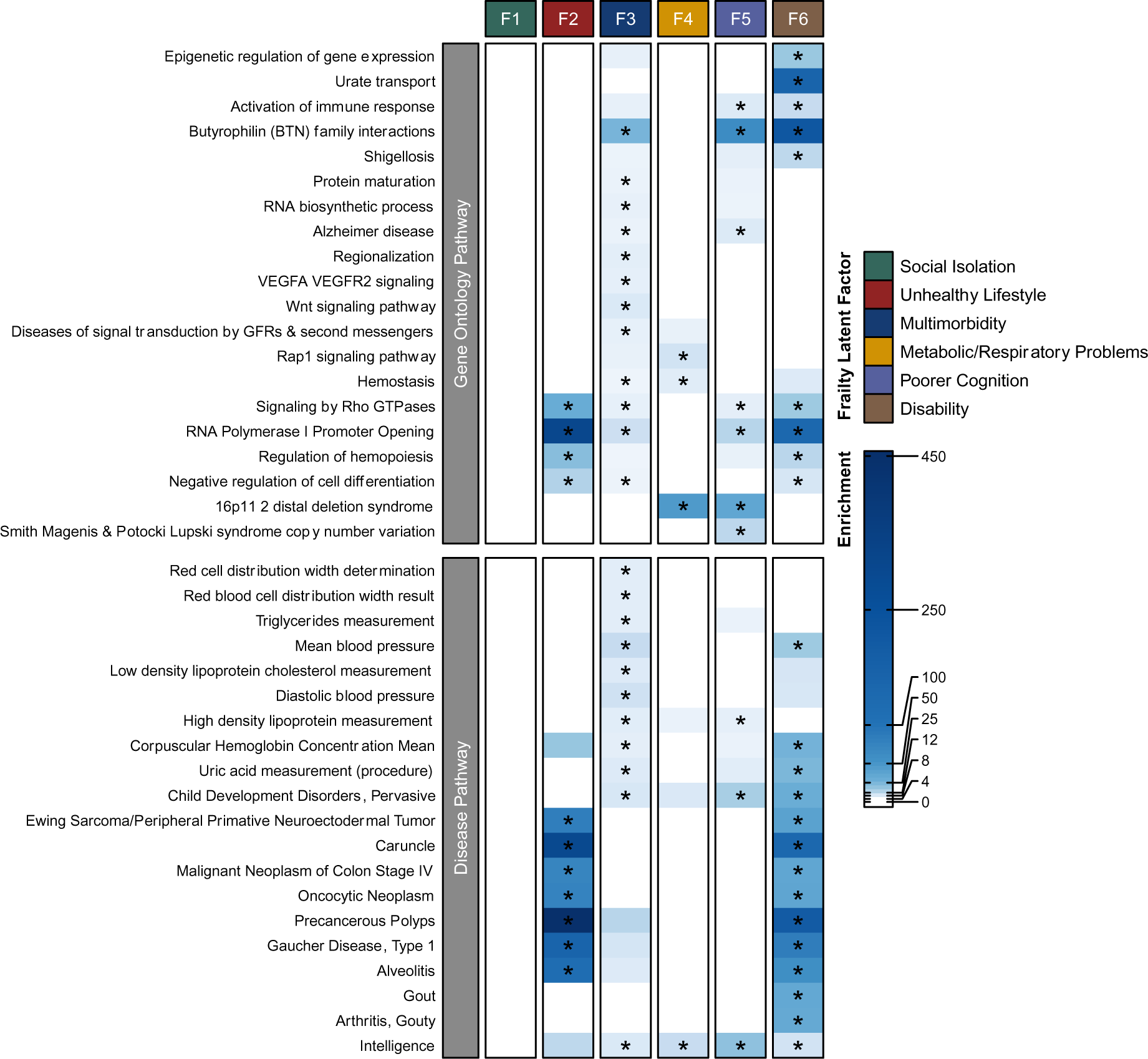
Results from pathway enrichment analysis of the prioritized genes for the residual frailty factors (Factors 1-6). The upper heatmap shows the enrichment for the top 20 enriched gene ontology pathway clusters. The displayed values represent the results for the most significant gene ontology term in the cluster (as named on the y-axis). The lower heatmap displays the enrichment for the top 20 most significantly enriched disease pathways from the DisGeNet database. There were no significantly enriched pathways for Factor 1 (social isolation) as only 4 genes (*CTNND1*, *TMX2*, *MED19* and *EGR3*) were mapped to that latent factor. Significant enrichment values are marked with an asterisk (*P_FDR_* < 0.05). GFRs = Growth Factor Receptors; VEGFA/R2 = vascular endothelial growth factor A/receptor 2; F1 = Factor 1; F2 = Factor 2; F3 = Factor 3; F4 = Factor 4; F5 = Factor 5; F6 = Factor 6.

Pathway analysis of the prioritized genes for the General Factor identified only two significantly enriched disease pathways for intelligence and scoliosis (**Tables S38-40).** In contrast, we found high levels of significant enrichment (i.e. *P_FDR_* < 0.05) for all residual latent frailty factors, except Factor 1 (**Figure 5** and **Tables S41-43**). The most strongly enriched pathway cluster (lead gene ontology term = RNA polymerase I promoter opening) included multiple gene sets linked to known aging-related processes including telomere function, amyloid fiber formation and oxidative stress, and was significantly enriched across Factors 2 (unhealthy lifestyle), 3 (multimorbidity), 5 (poorer cognition), and 6 (disability). Other pervasive cross-factor enrichment implicated immune function, epigenetic regulation, and cancer as key pathways involved in frailty pathogenesis. In addition to the shared enrichment in aging-related pathways observed across the factors, we also found evidence for the discriminant validity of the frailty factors. For example, only Factors 3 (multimorbidity) and 5 (poorer cognition) demonstrated significant enrichment for pathways linked to Alzheimer’s disease and general neurodegeneration. Factor 3 (multimorbidity) was also enriched in protein maturation and folding pathways, providing consistent evidence that aspects of frailty related to multimorbidity and cognition may be more highly linked to dementia and neurodegenerative pathways compared to other aspects of frailty. Factor 4 (metabolic/respiratory problems) genes were enriched in gene sets linked to cell signaling (particularly Rap1 pathways) and 16p11.2 distal deletion syndrome. This is a rare syndrome that results from the partial deletion of the short arm of chromosome 16 leading to symptoms including intellectual disability, developmental delay and autism spectrum disorder. This syndrome can be caused by unmasked recessive mutations in the *CLN3* gene(23), which was where the most significant risk locus for our frailty GWAS was located (lead SNP = rs27741; Factor 4 *P*-value = 1.09×10^−35^) (**Figure 3**). Enrichment analysis of disease pathways from the DisGeNet database further demonstrated that the frailty factors display distinct underlying biology. Factor 3 (multimorbidity) genes were strongly enriched in pathways linked to red blood cell and lipid biomarkers, whereas Factor 2 (unhealthy lifestyle) and 6 (disability) genes were significantly enriched in cancer pathways, and additionally, in gout and arthritis pathways for Factor 6.

### Polygenic Risk Scores of Frailty Factors Predict Frailty and Health in External Cohorts

To validate the latent frailty factors as phenotypes that capture frailty-specific variance, we created polygenic risk scores (PRSs) for each latent frailty factor and used regression models to test how well they predicted frailty and frailty-related outcomes in three external older adult cohorts (the Lothian Birth Cohort 1936 [LBC1936] (*N* = 1,005; mean age = 69.60), the English Longitudinal Study of Aging [ELSA] (*N =* 7,181, mean age = 68.45) and the Prospective Imaging Study of Aging [PISA] (*N* = 3,265, mean age = 60.34)) (**Online Methods** and **Supplementary Methods**). To measure the cumulative predictive capacity of our frailty model, we also created a PRS phenotype that combined the polygenic signal of all seven frailty factors using multiple regression (herein referred to as Multi-PRS). This allowed us to compare the performance of our overarching multivariate model in predicting frailty status relative to PRSs created from existing aggregate frailty GWAS measures (i.e. the FI-PRS(20) and FFS-PRS(21)). The combined Multi-PRS provided the strongest prediction of the FI across all three tested cohorts (**Table S44** and **Figure 6A-B**).

**Figure 6:**
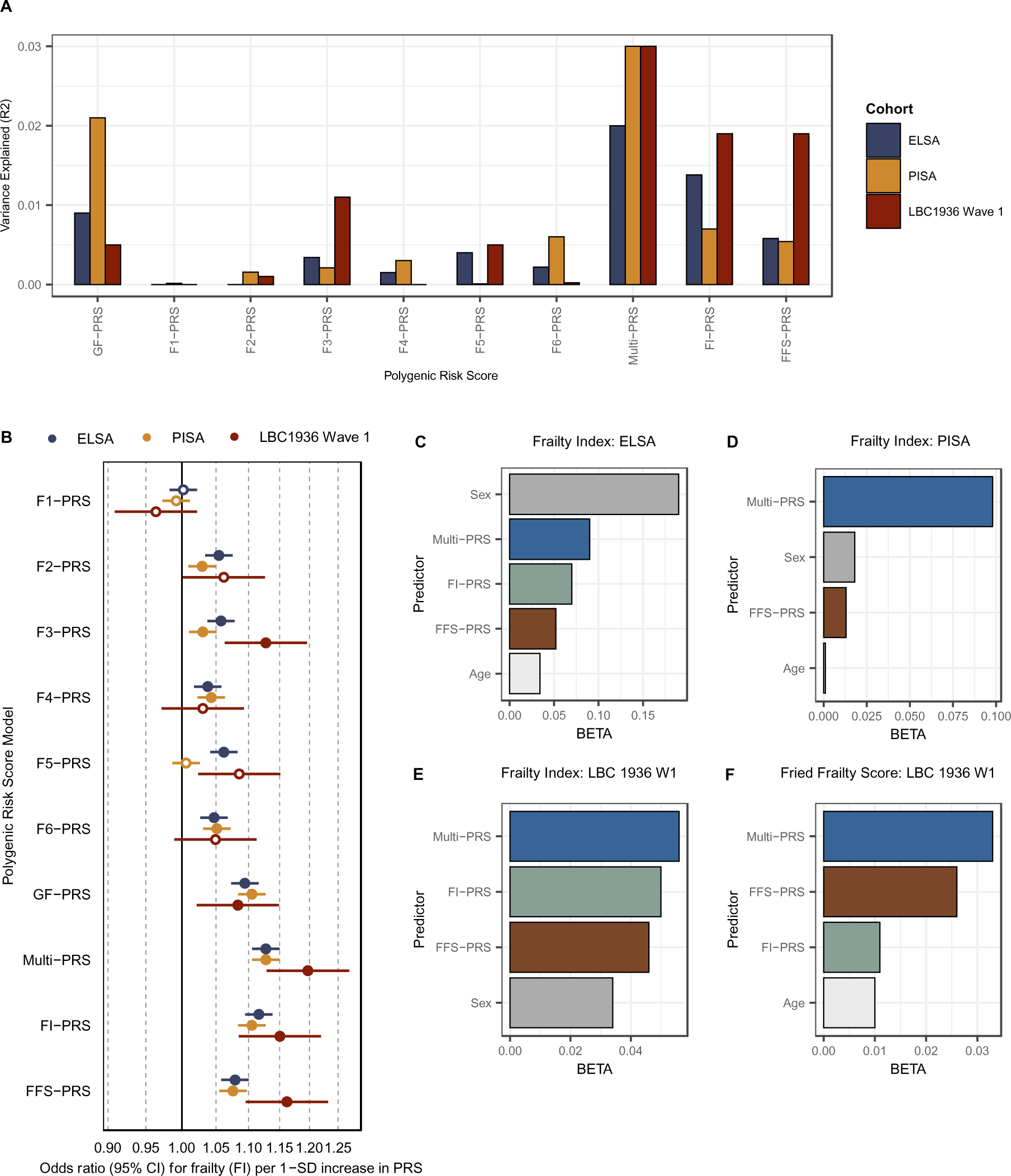
Results from the polygenic risk score (PRS) analysis of the latent frailty factors. *Panel A* displays a bar plot of the variance explained (R2) by each PRS that we estimated for prediction of the Frailty Index (FI) in each external cohort. *Panel B* depicts a forest plot of the odds ratios (95% Confidence Intervals [CI]) for each standard deviation (SD) increase in frailty (measured by the Frailty Index) associated with our frailty PRS phenotypes in each of the external cohorts. These values were calculated using linear regression models. Significant predictions (i.e. P_FDR_ < 0.05) are depicted as filled in dots, whereas non-significant predictions are depicted as an empty circle. *Panels C-F* represent the results from the elastic net regression analyses, which rank the performance of our Multi-PRS (i.e. combined latent frailty factor score) when modelled with the aggregate Frailty Index PRS (FI-PRS), the aggregate Fried Frailty Score PRS (FFS-PRS), age, sex and ancestral principal components as covariates. Only the predictors that explain significant additional unique variance over and above the other covariates are included in these graphs. All analyses represent standardized results. LBC1936 = Lothian Birth Cohort 1936; PISA = Prospective Imaging Study of Aging; ELSA = English Longitudinal Study of Aging; F1-F6 = Factors 1-6; GF = General Factor; W1 = Wave 1.

We additionally measured the association of the latent frailty PRSs with other frailty-related health outcomes, which helped to provide a more detailed picture of how these frailty subgroupings may differentially impact aging processes (**Table S45** and **Supplementary Methods**). We found that the PRS for Factor 5 (poorer cognition) and the Multi-PRS significantly predicted lower cognitive ability in LBC1936 (β = −0.65, SE = 0.10, *P_FDR_* = 1.43×10^−08^ and β = −0.62; SE = 0.10, *P_FDR_* = 3.34×10^−08^), but not cognitive change. The Multi-PRS was also significantly associated with dementia (β = 0.2, SE = 0.07, *P_FDR_* = 9.60×10^−03^) and stroke (β = 0.18, SE = 0.05, *P_FDR_* = 6.40×10^−04^) in ELSA.

We used elastic net regression to jointly model the PRS’s for the seven frailty factors so that we could rank the order that each contributed to predicting frailty status (**Supplementary Results** and **Table S46**). The General Factor of frailty was ranked as the highest contributor to FI prediction in ELSA and PISA. By contrast in LBC1936, the Factor 3 (multimorbidity) PRS was ranked highest in FI prediction while the General Factor PRS provided the most prediction of FFS status. We also used elastic net regression to rank the performance of the full latent frailty model (Multi-PRS) against the previously derived aggregate frailty GWAS measures (FI-PRS and FFS-PRS). We found that the Multi-PRS outperformed the FI-PRS and the FFS-PRS when predicting FI and FFS status in all tested datasets (**Figure 6C-F** and **Table S47**). Sensitivity analyses that grouped samples by age demonstrated that the predictive contribution of our Multi-PRS was stable across different age groups, whereas the contributions from the PRS of the individual latent frailty factors changed by age, suggesting that preventive or therapeutic interventions targeting specific frailty subgroups may be more successful if provided at specific points in the life course (**Supplementary Results**). Together, these findings validated our model as representing a novel genetic measure that captures frailty-relevant pathways, which explained more genetic variance than aggregate GWAS measures that have been used in the field so far. They also underline the importance of modeling subgroups within the frailty construct to better understand what drives frailty onset across the life course.

## Discussion

We report the first genomic factor analysis of frailty. We introduce seven novel latent constructs of the shared genetics between 30 frailty deficits, including a General Factor of frailty and six additional residual factors representing genetic overlap between distinct subsets of frailty deficits related to social isolation, unhealthy lifestyle, multimorbidity, metabolic/respiratory problems, poorer cognition, and disability. We identify 408 genomic risk loci for these latent constructs that are enriched for pathways related to accelerated aging, including epigenetic modifications and immune regulation. This demonstrates a substantial advance in genomic locus discovery for frailty compared to prior GWAS of aggregate frailty measures, which only identified 14 genomic loci for the FI(20) and 37 genomic loci for the FFS(21). We further validate the latent constructs as being of relevance to frailty and related health outcomes at multiple levels of biology and in the prediction of frailty status in external data.

Our findings support previous phenotypic studies that highlight the merit, relative to single aggregate scores, of using data reduction methods to improve our understanding of frailty etiology (12, 24, 25). However, by taking a multivariate genomic approach we were able to integrate theoretical knowledge with biological evidence to better define the underlying pathways of frailty and to differentiate generalized pathogenic pathways from more nuanced pathways that are specific to a subset of deficits, both of which are fundamental to understanding this complex clinical construct. For example, our genetic correlation and pathway analyses implicate immune function and epigenetic modifications as being key drivers of frailty pathogenesis across multiple deficit groupings. This is in line with findings linking frailty and elevated CRP levels, red blood cell distribution width, and white blood cell count (26–29). Our frailty factors were also significantly genetically correlated with health complications associated with infection, including hospitalization and sepsis. The associations between frailty and common viral infections, such as pneumonia(30), COVID-19(31) and urinary tract infections(32) are well documented. Furthermore, our findings consistently demonstrated evidence for widespread epigenetic changes in frailty, supporting previous work suggesting that epigenetic biomarkers, such as epigenetic clocks(33) or epigenetic risk scores(34) could be effective predictors of frailty.

The seven frailty factors displayed discriminant validity across multiple levels of biological analysis, indicating that existing aggregate measures of frailty are likely to miss clinically relevant distinctions. For example, we find that the poorer cognition factor (Factor 5) was uniquely associated with dementia pathways, particularly Alzheimer’s disease. In addition, our GWAS and gene prioritization findings implicated *SPI1* as a key locus for Factor 5, which is a well-replicated Alzheimer’s disease risk locus(35–37). Interestingly, Factor 5 had similar factor loadings from lower fluid intelligence and poor self-reported overall health rating, indicating that subjective health reports as well as cognitive testing could be indicative of subsequent heightened dementia risk in individuals who present with these frailty deficits. In fact, subjective cognitive decline has been widely supported as a potential early marker of cognitive impairment(38). The other loading onto this latent factor was slow walking pace, which is independently associated with heightened dementia risk(39). In addition, slow gait and subjective cognitive decline are used to measure motoric cognitive risk, a syndrome strongly associated with subsequent dementia(40).

Furthermore, our genetic correlation and PRS analyses found that the multimorbidity factor (defined by number of illnesses and high mean arterial pressure) is a strong driver of frailty over and above the variance captured by a general aggregate measure of all frailty deficits. Prevalence of multimorbidity and associated polypharmacy is a global public health concern, with rates as high as 90% in certain populations(41). This latent factor produced by far the highest number of genomic risk loci and PRS analyses demonstrated that its predictive power was substantial. Gene prioritization and pathway analysis indicated enrichment in a wide array of aging-related pathways, including *VEGFA* signaling, which was recently identified in a multivariate GWAS of aging(42) and has been shown to be important in longevity(43). Taken together, our findings suggest that this latent factor comprises a broad set of disease-related biological pathways that are associated with the most common diseases found within populations that lead to a heightened risk for developing frailty and accelerated aging. This provides empirical support for the ‘geroscience hypothesis’, which theorizes that manipulating aging physiology will prevent associated diseases (44).

Our findings should be viewed in light of several limitations. We did not explore the impact of sex differences, which are important in aging as evidenced by significant prevalence differences in frailty across all age groups(45). Our tissue enrichment analyses alluded to this with significant enrichment identified for the sex-specific reproductive organs. Future work should be performed to interrogate the sex-specific multivariate genetic architecture of frailty. Furthermore, our analyses were restricted to samples of European genetic ancestry since the methods rely on linkage disequilibrium information that can vary across ancestral populations. Unfortunately, despite advances in collecting genomic data from multiple populations, it was not possible to identify publicly available GWAS data for the frailty deficits to conduct a trans-ancestry analysis, but this should be a major focus in the future to make these results more generalizable globally.

In conclusion, we introduce the first genomic latent model of frailty. We demonstrate the added potential of modelling frailty as multiple latent factors, representing both a generalized pathway of frailty and distinct subgroups of deficits that share additional underlying biology. This can be contrasted to previous studies that have relied solely upon aggregate measures of frailty. This more nuanced model offered unique etiological insights into frailty and may aid in refining risk stratification of patients. Our genomic model of frailty may also help to develop novel preventive and therapeutic strategies that minimize the broad range of adverse frailty-related health outcomes.

## Online Methods

### Phenotype Selection

Phenotypes were selected based on deficits described by the FI(46). Phenotype selection was further guided by choosing traits that reflect systemic pathways and health behaviors (e.g. number of diagnosed illnesses) as opposed to specific clinical diagnoses (e.g. type 2 diabetes). This allowed for modeling genetic variance for general aspects of frailty rather than disease - specific pathways. Traits were included if they had well-powered GWAS summary statistics that were publicly available in a sample of ≥10,000 individuals of European ancestry. Analyses were restricted to European ancestry individuals as the methods used to estimate and model genetic overlap rely on ancestry-specific patterns of linkage disequilibrium (LD) and GWAS of sufficient sample size in other ancestry groups are not yet available across all the frailty traits. When possible, we prioritized the use of GWAS data from consortium-based studies since these tend to pool the largest sample sizes and have more rigorous phenotypic definitions (15–19, 47–52). When consortia data was not available, we used European ancestry GWAS summary statistics downloaded from the PanUKB (https://pan.ukbb.broadinstitute.org). **Table S1** displays a summary of the 52 traits included in our initial analysis. The effect estimates for each trait were formatted in a direction that reflected the ‘risk -inducing’ phenotype for frailty (i.e. slow walking pace as opposed to fast walking pace).

### Genetic Correlation Estimation

We conducted multivariable linkage disequilibrium (LD) score regression (LDSC) using the *GenomicSEM* R package along with publicly available LD scores and weights from the 1000 Genomes Phase 3 European reference panel that excluded the major histocompatibility complex (MHC) owing to its complex patterns of LD(53, 54). LDSC was applied to estimate a genetic covariance matrix with SNP-based heritability (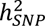) of each phenotype on the diagonal and the genetic covariance across each pairwise combination of traits on the off-diagonal. Multivariable LDSC also produces a sampling covariance matrix that includes the sampling variances (i.e., the squared standard errors) of the estimates on the diagonal and the sampling dependencies on the off diagonal that can arise due to participant sample overlap. This sampling covariance matrix is estimated directly from the GWAS data and is what allows Genomic SEM to produce appropriate estimates for traits with varying levels of precision (e.g., with different participant sample sizes) and varying and unknown levels of sample overlap. This was important for producing interpretable results within the current study, wherein our frailty deficits varied widely with respect to statistical power. Prior to running LDSC, each of the GWAS summary statistic datasets underwent a uniform formatting and quality control step using the *munge* function in the *GenomicSEM* R package. This included removing SNPs with a minor allele frequency (MAF) below 0.01 and an imputation score (INFO) below 0.9, restricting SNPs to HapMap 3 SNPs, and aligning all GWAS effects to the same reference allele (13). For all genome-wide analyses, we used a false discovery rate (FDR) corrected *p*-value threshold of <0.05 to account for multiple testing.

In line with the recommendations from the original LDSC developers, traits were removed from further analyses if they had a 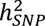 Z-statistic ≤4 as these can yield unstable estimates of genetic overlap(55). We converted these estimates to the liability scale for binary traits using population prevalence values based on recent epidemiological estimates for European ancestral populations (**Table S1**)(56–65). The sum of effective sample sizes across the cohorts that had contributed to that GWAS was used to adjust for cohort-specific ascertainment(66).

The results from the genetic correlation analysis (**Table S2**) were used to perform additional quality control, where deficits were brought forward for subsequent analysis providing they met the following criteria: (*i*) in instances where two traits were highly multicollinear (*r_g_* ≥0.9), we removed the trait in the pair that had the lowest mean genetic correlation (*r_g_*) with the other included traits; *(ii*) we removed traits that had low mean genetic correlations with the other measured traits (mean *r_g_* ≤0.10) as these traits are not empirically indicated to be part of the broader multivariate genetic architecture of frailty; (*iii*) for phenotypically similar traits (e.g., number of cigarettes smoked per day vs smoking initiation), we retained the deficit with the highest mean *r_g_* with all other deficits. This resulted in a final list of 30 traits that were brought forward for all subsequent analyses (**Table S1**). A frailty index constructed with 30 or more deficits has been shown to sufficiently capture frailty in phenotypic literature(8).

### Genomic Factor Analysis

#### Exploratory Factor Analysis

Since the latent pattern of the shared genetic architecture between frailty deficits has not been assessed previously, we initially ran an exploratory factor analysis (EFA) using the *stats* R package to identify a plausible latent structure that describes the genetic overlap across the included frailty deficits. To avoid model overfitting, we used the genetic covariance matrix estimated in odd autosome as the input to the EFA and the genetic covariance matrix estimated in even autosomes as the input for the subsequent confirmatory factor analysis (CFA). We used the Kaiser rule(67) and the number of optimal coordinates test(68) to determine the number of factors to extract in the EFA, which both suggested that seven factors were appropriate. We additionally extracted a 6-factor model (**Table S48**) as there were a high number of cross-loadings in the 7-factor specification, indicating a more parsimonious structure may be appropriate, and the 5^th^ factor in the 7-factor model only captured genetic variance related to mean arterial pressure (**Table S49**). Owing to highly pervasive correlations between our frailty deficits (**Table S2**), we applied promax factor rotation, which allows for inter-factor correlations.

#### Confirmatory Factor Analysis

We then conducted a CFA using the genetic covariance matrix from the even autosomes as input. The CFA model was guided using the EFA in odd autosomes, where a frailty deficit was specified to load on a factor when standardized loadings were ≥0.30. Diagonally weighted least squares (DWLS) was used for model estimation and any frailty deficits that had negative residual variances were constrained to have a residual variance >0.001. The 6-factor (Comparative Fit Index [CFI] = 0.92; Standardized Root Mean Square Residual [SRMR] = 0.07) and 7-factor model specification (CFI= 0.89; SRMR= 0.07) both provided good fit to the even autosome data (**Table S50**). The 6-factor model was selected over the 7-factor model as it: (*i*) provided improved fit to the data while offering a more parsimonious representation of the data relative to the 7 -factor structure; (*ii*) it produced more theoretically interpretable factors of latent genetic architecture between distinct groups of multiple frailty deficits compared to the 7-factor model; and (*iii*) the 6-factor model continued to provide good fit to the data in all autosomes (CFI= 0.92; SRMR = 0.06) (**Table S51**).

#### Bifactor Model

While the 6-factor model produced theoretically meaningful latent factors, the first latent factor displayed strong factor loadings for 16 out of the 30 frailty deficits and the model included pervasive cross-loadings of frailty deficits on multiple factors. Taken together, these findings indicated that a bifactor model was an appropriate way to capture the general frailty pathways across all included deficits, as well as the genetic variance specific to distinct subsets of deficits.

Therefore, we estimated the fit of a bifactor model that included loadings for all 30 frailty deficits onto a general factor of frailty (herein termed General Factor), in addition to the loadings on the 6 latent factors from the CFA model (herein named Factors 1-6). A key benefit to this approach is that the General Factor is orthogonal (i.e. uncorrelated) to the additional residual group factors, which enabled us to interpret the General Factor as general genetic pathways of frailty that are distinct from the more focused subsets of genetic variance that underlie potential subgroups within the frailty spectrum. A bifactor model thereby provided a more direct test of our hypothesis that aggregate scores of frailty (e.g., the FI and FFS) miss unique risk pathways that are only shared between smaller subsets of frailty deficits.

Owing to the inclusion of the General Factor, some of the original factor loadings for the six CFA factors became nonsignificant. We iteratively removed any loadings from Factors 1-6 that were below our 0.30 threshold to ensure that we only retained stable loadings in the final model specification. In cases where an indicator displayed loadings above our cut-off for multiple residual factors, we retained these cross-loadings because a previous simulation study found that omitting substantial cross-loadings from a bifactor model based on a prior CFA model can upwardly bias the general factor loadings and downwardly bias residual group factor loadings, which cannot be picked up using standard model fit measures(69).

We allowed the residual group factors (Factors 1-6) to be correlated (but orthogonal to the General Factor). This form of bifactor model is known as a bifactor(S-1) model and is sufficiently identified if a subset of the indicators only load onto the bifactor (in our case 50% of the frailty deficits solely loaded onto the bifactor)(70). To ensure the model was locally identified, factor loadings were constrained to be equal when there were only two indicators that loaded onto a factor (i.e., for Factors 1 and 3). The final bifactor(S-1) model (**Figure 1** and **Table S3**) continued to provide good fit to the data (CFI = 0.93; SRMR = 0.07) and was brought forward for all subsequent analyses.

### Genetic Correlations with Related Health Traits

Frailty is known to increase the risk of many adverse health outcomes, but it is unclear whether this is owing to shared genetics between the more general frailty pathways or whether some outcomes are only associated with certain deficits within the frailty state. Furthermore, since this represented the first time that frailty has been measured in this latent framework, we wanted to validate our factors as reflecting frailty since well-powered independent GWAS samples for all 30 deficits are not currently available to do a full replication GWAS. Therefore, we performed genetic correlation analyses to assess the associations between 52 aging-related health outcomes and frailty phenotypes to assess the different patterns of shared genetics between these outcomes and each of the latent factors (**Table S4-S5**)(20, 21, 71–90). We used the same quality control procedures and data curation steps on the GWAS summary statistics for these 52 health outcomes as described for the main frailty deficits using the *munge* function in the *GenomicSEM* R package. However, in the case of pneumonia there were no prior available GWAS summary statistics with a SNP-based heritability estimate high enough to be included in LDSC. Therefore, we conducted a fixed-effect meta-analysis using METAL software, which comprised publicly available GWAS summary statistics data from the European ancestry sample in PanUKB (https://pan.ukbb.broadinstitute.org; N_Cases_ = 14,054 and N_Controls_ = 405,999) and FinnGen release 10 (https://r10.finngen.fi; N_Cases_ = 63,377 and N_Controls_ = 348,804). This resulted in a total GWAS sample of 832,234 individuals (77,431 pneumonia cases and 754,803 healthy controls), which produced a reasonable SNP-based heritability *Z*-statistic for LDSC (**Table S4**; *Z*-statistic = 4.1). We then constructed a separate genome-wide genetic variance and covariance matrix for each of the 52 external traits combined with the 30 frailty deficits using multivariable LD score regression. We re-specified our latent frailty model with an additional correlation between the external trait and each latent factor to get the genetic correlation and standard error estimates for each of the external traits with each latent factor. We used an FDR-corrected *p*-value threshold <0.05 to correct for multiple testing.

### Multivariate GWAS of Latent Frailty Factors

#### Data Standardization

We performed a multivariate GWAS within the *GenomicSEM* R package that estimated the individual SNP associations with each of the latent factors in our bifactor(S-1) model. We initially used the *sumstats* function to create a standardized data file that converted the SNP effect estimates from each univariate GWAS into covariances between each individual SNP and the overall phenotypic variance of each deficit and standardized the SNP effects relative to the phenotypic variance of the trait (13). This allowed these SNP effects to be added to the genetic covariance matrix as they were then on the same scale as the LDSC estimates. All phenotypes were based on genome build 37 (GRCh37/hg19) or were converted to build 37 using the *MungeSumstats* R package(91). We removed any SNPs that had a MAF <0.01 or an imputation score (INFO) <0.6 according to recommended defaults within the package. We used the 1000 Genomes Phase 3 European ancestry dataset as our reference genome and aligned SNPs to have the same effect (A1) and non-effect (A2) allele across phenotypes. SNPs were removed if either the effect or non-effect allele did not match the reference genome. As *sumstats* performs listwise deletion across the included traits only SNPs that had been measured in all the contributing univariate GWASs were included, resulting in a final harmonized dataset of 5,849,452 SNPs for multivariate GWAS analysis.

#### Multivariate GWAS Estimation

The standardized GWAS data and LDSC results were used as input for calculating multivariate GWAS of the frailty latent factors using the *userGWAS* function in the *GenomicSEM* R package. We fixed the measurement model (i.e. the genome-wide factor loadings and factor correlations) for all SNPs. This improved computational tractability and model interpretability as the SNP-specific estimates were scaled according to the same measurement model across all SNPs (as opposed to the entire model being re-estimated for each SNP). We removed any SNPs that required a high level of smoothing (i.e. *Z*-statistic change pre- and post-smoothing >1.96) or that produced lavaan warnings for negative observed variable or latent variable variances or non-positive definite covariance matrices (293 SNPs removed in total).

#### Q_SNP_ Heterogeneity Index

As previously described, not all the genetic signal captured within the latent factor GWAS results represent genuinely shared genetic variance. For example, strong signal from a single indicator (i.e. the *FTO* locus for body fat percentage) can lead to false positives if not properly accounted for(13). Likewise, some of the non-significant genetic signal within the multivariate GWAS results may represent areas of the genome that have highly heterogenous magnitudes and directions of effects on the different univariate indicators(13). For this reason, it is necessary to calculate the Q_SNP_ heterogeneity statistics for each SNP, which reflects a χ^2^ distributed statistic that is more significant for SNPs whose effects deviate strongly from the patterning of effects implied by the factor model. As part of the current project, we introduce and validate a more computationally efficient way of calculating Q_SNP_. While the prior formulation of Q_SNP_ required estimating a series of follow-up models to calculate the heterogeneity statistic, our new formulation is automatically calculated for each factor predicted by a SNP in the model. This change thereby greatly reduced the run-time of our analysis. The new Q_SNP_ equation starts by calculating the residual covariance matrix for the subset of the matrix that reflects the SNP-phenotype covariances for the phenotypes that load on a given factor (RSNP) as:

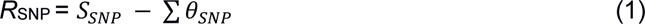

where *S*_*SNP*_ is the vector of SNP-phenotype covariances and ∑ *θ*_*SNP*_ reflects the model implied SNP-phenotype covariances. These model implied estimates reflect the product of the estimated SNP-effect on a given factor and the factor loadings for each trait. The precision of those SNP-phenotype estimates is indexed by taking the eigen decomposition of the portion of the sampling covariance matrix (*V*) that indexes those SNP-phenotype effects (*V_SNP_*):

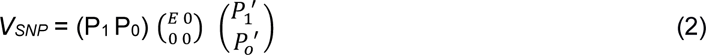

where *P_1_* is the matrix of principal components (eigenvectors) of *V_SNP_*, *P_0_* is the null space of *V_SNP_*, and *E* is a diagonal matrix of the non-zero eigenvalues of *V_SNP_*. These eigenvalues and eigenvectors can then be used to weight the residual covariance matrix of the SNP-phenotype estimates to obtain a *χ*^2^ distributed test statistic given as:

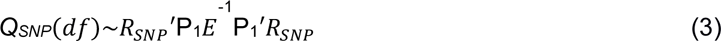

where *df* reflects the degrees of freedom, which will be one less than the number of indicators for the factor. We note that this equation is iteratively applied for each factor that is predicted by a SNP, such that a separate *R*_SNP_, *V_SNP_*, and factor-specific *Q_SNP_* are produced.

We demonstrate via simulation that this new approach continues to produce a *χ*^2^ distributed statistic that is statistically equivalent to the previously described formulation of Q_SNP_. We used the *simulateData* function within the *lavaan* R package to simulate data for 3 different factor models each with 50,000 observations for 1000 SNPs. We tested a 2 -factor model with 3 indicators on each factor (2 degrees of freedom [df]), a 2 -factor model with 4 indicators on each factor (3 df) and a 2-factor model with 6 indicators on each factor (5 df). We confirmed across all 3 examples that the new method remained *χ*^2^-distributed and that they did not differ significantly from the estimates calculated using the old method in terms of the mean Q_SNP_ (**Supplementary Results**). In addition, the new method consistently demonstrated a well-calibrated type 1 error rate (*p* < 0.05) (**Supplementary Results**).

For our empirical frailty application, we pruned out the Q_SNP_ significant signal from our GWAS summary statistics for each latent frailty factor to ensure that we only measured shared genetic variance operating via each latent factor in our subsequent post -GWAS analyses. We did this by removing SNPs that had a Bonferroni-corrected Q_SNP_ p-value <7.14×10^−09^ (i.e. 5×10^−08^/7) and any SNPs that were within a 1 megabase window upstream or downstream of this location to ensure that variants that were in LD with these heterogenous regions were removed.

Once the Q_SNP_ signal had been pruned from the latent factor summary statistics, we used the method developed by Mallard and colleagues to calculate the expected sample size (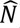) of each latent factor(92) using the following equation:

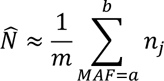

Where the expected sample size (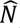) is approximately equal to the mean SNP expected sample size (*n*_*j*_) for *m* number of SNPs that have a MAF between *a* and *b* (in this case we used a MAF range of SNPs between 10-40% as these provide more stable estimates)(92). This value quantifies the amount of error-free genetic variance that is being captured by each latent factor, so this value can also act as an indicator of how well-powered each latent factor is within the model.

#### Identification of Genomic Risk Loci for Latent Frailty Factors

We used FUMA v1.5.2 to identify genomic risk loci for each latent factor in our model using the default software parameters (93). We used a Bonferroni corrected genome-wide significance threshold of *p* < 7.14×10^−09^ (i.e. 5×10^−08^/7 factors) to identify significant SNPs within our pruned GWAS summary statistics for each latent factor (i.e. Q_SNP_ significant variants removed). A genomic risk locus was defined as the region around a genome-wide significant SNP that included all SNPs that were in LD (*R*^2^ < 0.6) with that variant based on LD patterns in the 1000 Genomes Phase 3 European ancestry reference genome(93). If there were additional independently significant SNPs in LD with the lead SNP (*R*^2^ > 0.1) or if loci were located within 250 kilobases (kb) of one another, these were merged into a single locus(93).

### Gene Mapping and Functional Annotation

As an initial step to understand the biological implications of the loci underlying each latent frailty factor, we applied the SNP2GENE function in FUMA v1.5.2 to map potentially causal genes to each locus and to functionally annotate the candidate SNPs by integrating our results with various external datasets(93). This involved positionally mapping candidate SNPs to their closest protein-coding gene (within a 10kb window upstream or downstream of the SNP location). We conducted expression quantitative trait loci (eQTL) mapping to ascertain whether the candidate SNPs in our latent factor GWAS loci are known regulators of gene expression, which we defined as any eQTL-gene interaction displaying a *P_FDR_* ≤ 0.05(93). Since frailty represents a multi-system clinical state, we integrated publicly available eQTL data for multiple body tissues and cell types with our GWAS results to provide a broad overview of the potential impacts that the risk loci for frailty have on gene expression. This included integration of eQTL data from 15 repositories including the Blood eQTL Browser(94), BIOS QTL browser(95), BRAINEAC(96), MuTHER(97), xQTLServer(98), CommonMind Consortium(99), eQTLGen(100), PsychENCODE(101), DICE(102), scRNA eQTLs(103), eQTL Catalogue(104–120), GTEx v8(121), EyeGEx(122), InsPIRE(123) and TIGER(124). Finally, we conducted chromatin interaction mapping, which involved annotating candidate SNPs that displayed a significant 3D chromatin interaction (*P*_FDR_ ≤ 1×10^−6^) with a known gene region based on pre-processed chromatin loop data and enhancer-promoter link data for multiple body tissues from 4 publicly available databases(101, 125–127). We restricted successful mappings of SNPs to those where the candidate SNP overlapped with the enhancer region within one of the selected epigenomes and the promoter region of the mapped gene overlapped with the promoter region of the associated epigenome. Enhancer and promotor information was obtained for 111 epigenomes from the Roadmap Epigenomics Project(128).

We additionally used MAGMA v1.08(129) to conduct a gene-based analysis that identified genes that were significantly associated with each latent factor. Any SNPs that were present in the GWAS summary statistics for the latent factor and that were located within one of the protein-coding genes in the Ensembl database (excluding the MHC region) were analyzed. We used the 1000 Genomes Phase 3 European dataset as our LD reference panel and applied a Bonferroni correction for number of genes tested within each latent factor GWAS(93). Using gene expression data from GTEx v8 for 54 body tissues(121), we also used MAGMA to perform a gene property analysis to ascertain whether the genes that were significantly associated with the latent factor in the gene-based test were more likely to produce gene expression changes in particular body tissues(130).

### Stratified Genomic SEM

Since frailty has been consistently linked to increased risk of poorer brain health and dementia, which represents a key burden on health services in aging populations(8), we explored whether there was evidence for brain-relevant functional enrichment in the genetic variance captured by our latent frailty factors. We applied Stratified Genomic SEM(56) to test whether there was evidence for enrichment in functional annotations (groups of genetic variants combined due to having a shared biological characteristic) that are known to influence tissue-specific gene expression in different brain regions, histone modifications, neuronal cell types, or the interaction between these neuronal cell types and protein-truncating-variant-intolerant (PI) genes. We used previously constructed functional annotations based on data from 1000 Genomes Phase 3 Baseline LD Version 2.2(131), GTEx(132), DEPICT(133), the Roadmap Epigenetics Project(128) and the Genome Aggregation Database (gnomAD)(134), which consisted of a total of 172 functional annotations. This included 5 randomly selected non-brain control regions for the gene expression and histone modifications(135).

We performed multivariable stratified LDSC to estimate the zero-order genetic covariance matrices and the corresponding sampling covariance matrices that were partitioned across the genomic regions of each functional annotation using the *s_ldsc* function in the *GenomicSEM* R package(56, 131). We subsequently used these matrices as the input data to the *enrich* function to calculate an enrichment ratio for the genetic variance captured within each latent factor in our bifactor model for each functional annotation(136). We removed 26 functional annotations from our analyses owing to high degrees of smoothing (defined as a Z-score difference >1.96 pre and post-smoothing), as this indicates low power to detect meaningful enrichment(135). This resulted in 146 functional annotations being retained in our analyses. We used an FDR corrected *p*-value threshold of <0.05 to account for multiple testing.

### Summary Data-Based Mendelian Randomization

We applied multi-SNP summary data-based Mendelian randomization (SMR-multi) to prioritize genes for each latent frailty factor by identifying SNP-outcome (i.e. variant-frailty) associations that demonstrated strong evidence for being driven via intermediary pleiotropic effects on gene expression (eQTLs), splicing ratios (sQTLs) or methylation status (mQTLs; (137, 138). We used pre-processed data curated by the original method developers, which included eQTL data from BrainMeta v2(139), CAGE(140), GTEx v8(121), GEUVADIS(111), PsychENCODE (corrected for 100 hidden covariate factors)(141) and Westra et al(94); sQTL data from BrainMeta v2(139) and GTEx v8(121); and mQTL from Brain-mMeta(142) and McRae et al(138, 143). All data was based on genome build 37 (GRCh37/hg19) and we aligned the GWAS data and QTL data to the same effect and non-effect alleles using the 1000 Genomes phase 3 European data as our reference. We converted the MAF column in the GWAS summary statistics into the effect allele frequency (EAF) by integrating data on the major allele for each SNP from dbSNP v155. Any SNPs that had a discrepant allele frequency >0.20 between the GWAS and QTL data or that were located within the extended MHC region (chr6:28477797-chr6:33448354) were omitted from the analysis(138). SNPs that had a strong association (*P*QTL < 5×10^−8^) with the QTL region (i.e. expression, splicing or methylation) in each QTL dataset were selected as instrumental variables for SMR-multi analysis(138). Variants in high LD with the lead SNP in a region (*R*^2^ > 0.9) were removed using the 1000 Genomes phase 3 European data as a reference. We then applied FDR correction across all the results (i.e. all tests conducted across all latent factors and all QTL datasets) and filtered our results to only include SMR associations with *P*SMR-FDR < 0.05. To distinguish pleiotropy (i.e., association between exposure and outcome due to shared causal variant) from linkage (i.e., association between exposure and outcome due to independent causal SNPs being in LD with one another) we used the heterogeneity in dependent instruments (HEIDI) test using the default parameters to measure whether the association patterns within an instrumental variable were homogenous or not, and we rejected any associations that demonstrated significant heterogeneity (*P*_HEIDI_ < 0.01)(138).

### Gene Prioritization and Pathway Analysis

In order to better understand the underlying biological pathways of each latent frailty factor, we conducted pathway analysis using the genes that had been prioritized by the aforementioned gene mapping analyses as input. However, since not all genes that are mapped to risk loci represent truly causal genes, we triangulated our results to only include genes that presented sustained evidence for being a potential causal candidate for each latent factor. We defined this as any gene that was mapped by ≥ 3 of our gene mapping methods (i.e. positional mapping, eQTL mapping, chromatin interaction mapping, MAGMA or SMR-multi). We used METASCAPE (22) to perform a pathway enrichment analysis to identify gene sets that were significantly overrepresented in the prioritized genes for each latent factor. Since the General Factor was orthogonal to the six latent residual group factors in our model, we conducted a single standalone pathway enrichment analysis for the General Factor prioritized genes because, by definition, the genetics underpinning this latent factor are uncorrelated with the genetics driving the other latent factors. We then performed a separate combined enrichment analysis of the prioritized genes for Factors 1-6 as this enabled us to explore what biology might be driving the inter-factor correlations that we saw in our model. The prioritized genes were assigned to any curated gene set they belonged to using data from Gene Ontology (GO) Biological Processes (1495 gene sets)(144), the Human Molecular Signatures Database (MSigDB) Canonical Pathways (17 gene sets)(145), Reactome (262 gene sets)(146), Kyoto Encyclopedia of Genes and Genomes (KEGG) Pathways (113 gene sets)(147), WikiPathways (89 gene sets)(148) and DisGeNET (2050 gene sets)(149). The default hypergeometric test method was used to calculate enrichment for each gene set that included ≥ 1 of the prioritized genes and FDR correction was applied to correct for multiple testing(22). However, many of the gene ontology terms overlap, which leads to high levels of redundancy in the results, so we performed the recommended clustering analysis that combined related gene sets into groups by calculating the pairwise similarities between all enriched terms based on their Kappa-test score and hierarchally ordered them within a similarities matrix (Kappa similarities > 0.3 were combined)(22). In the case of the multi-gene-list enrichment test for the analysis of Factors 1-6, this enrichment test was initially performed on each of the latent factor gene lists separately, followed by an additional test that combined the gene lists across all six latent factors to determine whether there were enriched gene ontology terms that were shared across the latent factors. This was appropriate since our initial results demonstrated that there was significant correlation between certain latent factors in our model.

### Polygenic Risk Scores

#### Polygenic Risk Score Construction

To externally validate our frailty latent factors, we constructed polygenic risk scores (PRS) of each latent frailty factor and tested whether they predicted frailty and related health outcomes in three external cohort datasets, including the Lothian Birth Cohort 1936 (LBC1936), the English Longitudinal Study of Ageing (ELSA), and the Prospective Imaging Study of Ageing (PISA) (see **Supplementary Methods** for sample descriptions). We used the GWAS summary statistics of the shared genetic signal for each of the latent frailty factors (i.e., the summary statistics that had removed significantly heterogeneous signal), as well as publicly available GWAS summary statistics (downloaded from GWAS Catalog(150)) from previously published studies of aggregate measures for the FI(20) and FFS(21) to construct a separate PRS for each of these predictor phenotypes. This enabled us to compare the prediction of the latent frailty factors with routinely used aggregate frailty measures. We performed routine quality control on each of the base datasets (i.e. the GWAS summary statistics) and the target datasets (i.e. individual-level cohort genetic data). We aligned the effect and non-effect alleles across all datasets to ensure that the direction of effect was concordant across analyses. We removed SNPs with a MAF < 0.01, as well as duplicate and ambiguous SNPs.

Following quality control, PRS were calculated for the individuals in each of the three cohorts using PRSice-2 for LBC1936 and ELSA, and SBayesR for the PISA cohort. We followed default procedures for each methodology that have been described in detail by the respective developers(151, 152). Briefly, PRSice-2 uses a *P*-value based clumping and thresholding (P+T) approach in which the GWAS effect size estimates are used as SNP weights in the cohort data and clumping is applied to remove SNPs in high LD so that the final PRS only includes independent significant SNPs(151). In contrast, SBayesR is a Bayesian-based method, which estimates joint SNP effects across the genome using multiple linear regression whilst assuming a finite mixture of normally distributed priors(152).

#### Prediction of Frailty and Related Phenotypes in the External Cohorts

We subsequently performed a series of analyses to explore how well the latent frailty factors predicted routinely measured frailty phenotypes and related traits in external data. First, we used linear regression models to assess how well each individual frailty latent factor PRS predicted the FI in LBC1936 (based on 30 deficits), ELSA (based on 62 deficits) and PISA (based on 69 deficits) and logistic regression models to measure how well they each predicted the FFS in LBC1936 (see **Supplementary Methods and Tables S52-54** for details on the outcomes used). Since the latent frailty factors each represented distinct genetic variance that can contribute to frailty, we also used multiple regression to calculate a PRS of the combined scores for all 7 latent factors (i.e. Multi-PRS). Finally, to enable us to compare the performance of the latent factor PRS with previously published frailty GWAS aggregate measures, we also tested how well the FI GWAS PRS (FI-PRS) and the FFS GWAS PRS (FFS-PRS) predicted the same frailty outcomes in each dataset. All models included age, sex and ancestry principal components ([PCs]; for ELSA and PISA we used 10 PCs and for LBC1936 we used 4 PCs) as covariates. These models allowed us to calculate the amount of incremental phenotypic variance explained (*R^2^*) by each PRS, which was calculated by subtracting the covariate-only model *R^2^* from the *R^2^* of the full PRS and covariate model(153).

To assess the association of the latent factor PRS’s with frailty-related health outcomes, we also conducted regression analyses to test the association of each frailty PRS with cognitive ability (LBC1936), cognitive change (LBC1936), dementia (ELSA), motoric cognitive risk (LBC1936), mortality (LBC1936), stroke (LBC1936, ELSA and PISA) and memory complaints (PISA) (see **Supplementary Methods** for phenotype and analysis descriptions).

#### Elastic Net Regression to Rank Performance of Frailty PRS

Finally, since conventional linear regression models can be upwardly biased due to model overfitting, we performed elastic net regularized regression models in all three cohorts to rank the polygenic contributions to frailty whilst minimizing bias from model overfitting and multicollinearity between predictors(154). This method allows highly genetically correlated variables to be grouped and the final coefficients returned in the model allow the predictors to be ranked by their contribution of prediction to the outcome(154). Therefore, we ran an initial model predicting the FI in each cohort using the seven individual latent frailty factor PRSs and covariates (age, sex and ancestry PCs) as predictors to rank the latent factors in order of their strength in predicting frailty. We then performed elastic net regression that included the Multi-PRS, FI-PRS and FFS-PRS and covariates (age, sex, and 10 ancestry PCs) to rank prediction of the different genetic measures of frailty (i.e. multiple latent factors versus aggregate measures for the FI and FFS).

## Data Availability

The latent frailty factor and pneumonia GWAS summary statistics that were created by this study will be made publicly available on <u>GWAS Catalog</u> upon publication. All of the GWAS summary statistics that were used in this study for the Genomic SEM and genetic correlation analyses are publicly available via the original articles or from <u>PanUKB</u> and <u>FinnGen</u>.

## Code Availability

This code was developed using publicly available software that is available via the following links:

Genomic SEM (including our new Q_SNP_ extension): https://github.com/GenomicSEM/GenomicSEM

METAL: https://genome.sph.umich.edu/wiki/METAL

FUMA: https://fuma.ctglab.nl

SMR: https://yanglab.westlake.edu.cn/software/smr/

METASCAPE: https://metascape.org/gp/index.html#/main/step1

SBayesR: https://cnsgenomics.com/software/gctb/#Overview

PRSice-2: https://choishingwan.github.io/PRSice/

Lavaan: https://lavaan.ugent.be

MungeSumstats: https://github.com/neurogenomics/MungeSumstats

The specific code for the analyses in this study will be made available at https://github.com/IsyFoote upon publication. The code we used to create the latent growth curve models of cognitive ability and cognitive change in LBC1936 can be found here: https://lothianbirthcohorts.github.io/longitudinal-g-models/longitudinal_g_models

## Author Contributions

IFF, ADG, JDF, AR, JMR, DJL, TKK and KR conceived the study. IFF, JPF and AEF performed the analyses. SRC, ML, NGM and MKL provided key data access and support for the study analyses. JPF and DSM performed the data curation of phenotypes in the external individual-level datasets. IFF, ADG and JPF wrote the initial manuscript. All authors contributed to the design and conceptual aspects of the study and critically reviewed the final manuscript.

## Acknowledgements

This study was facilitated by the Deep Dementia Phenotyping (DEMON) Network through the Frailty and Dementia Special Interest Group and is an outcome of a workshop entitled “Frailty and Precision Dementia Medicine” funded by the University of Exeter Global Partnerships Fund (JMR). This work was funded by grants from the National Institute on Aging (IFF, AEF and ADG; RF1AG073593) and the National Institute of Mental Health (ADG; R01MH120219). The DEMON Network is supported by Alzheimer’s Research UK (DJL and JMR). DJL is also supported by the National Institute for Health and Care Research Applied Research Collaboration South West Peninsula. The Lothian Birth Cohorts 1936 (LBC 1936) is supported by the Biotechnology and Biological Sciences Research Council, the Economic and Social Research Council [BB/W008793/1], Age UK (The Disconnected Mind Project), the Milton Damerel Trust, and The University of Edinburgh. SRC is supported by a Sir Henry Dale Fellowship jointly funded by the Wellcome Trust and the Royal Society (221890/Z/20/Z). The English Longitudinal Study of Aging (ELSA) is funded by the National Institute on Aging (R01AG017644), and by UK Government Departments coordinated by the National Institute for Health and Care Research (NIHR). The Prospective Imaging Study of Ageing (PISA) is funded by the National Health and Medical Research Council (Grant ID: APP1095227). JPF is funded by the Legal & General Group (research grant to establish the independent Advanced Care Research Centre at the University of Edinburgh) - legal & General had no role in the conduct of the study, interpretation, or the decision to submit for publication - the views expressed are those of the authors and not necessarily those of legal and general. JDF receives research grant support from the Canadian Institutes of Health Research, the National Multiple Sclerosis Society, MS Canada, and consultation and distribution royalties from MAPI Research Trust. ADR thanks the Natural Sciences and Engineering Research Council of Canada (NSERC) for operating grant RGPIN-2019-05888. KR is supported by Dalhousie University Faculty of Medicine Advancement funding as Clinical Research Professor of Frailty and Aging.

We would like to thank the authors of the original GWAS publications that we used in this study for making their data publicly available, including the Neale lab and FinnGen (for unpublished by freely available GWAS data from UK Biobank and FinnGen). We would also like to thank the participants of the contributing cohorts and the research teams for providing their data for this work.

