## Supplementary Materials for "Uncovering the multivariate genetic architecture of frailty with genomic structural equation modelling"

### Supplementary Methods

#### Description of the Cohort Samples Used for the Main Polygenic Risk Score Analyses

***Lothian Birth Cohort 1936 (LBC1936).*** LBC1936 is a longitudinal study of community dwelling older adults living in and around Edinburgh, Scotland (United Kingdom)(1). Individuals joined the study when they initially took part in the Scottish Mental Survey in 1947 and have thus far taken part in 5 waves of testing. The main analyses, when examining frailty outcomes, were performed on wave 1 (mean age = 69.60; SD = 0.83; *N* = 1005 (509 males, 496 females)). Ethics for LBC1936 was approved by the Multi-Centre Research Ethics Committee for Scotland (Wave 1: MREC/01/0/56), the Lothian Research Ethics Committee (Wave 1: LREC/2003/2/29), and the Scotland A Research Ethics Committee (Waves 2, 3, 4 and 5: 07/MRE00/58) and all methods were performed in accordance with the relevant guidelines and regulations. Informed Written Consent was obtained from participants at each of the waves. The genotypes, which were collected via blood samples from the participants at Wave 1 were processed using stringent quality control measures(2).

***English Longitudinal Study of Aging (ELSA).*** ELSA is a longitudinal panel study of community dwelling adults aged 50 years and over residing in England (United Kingdom)(3). For the main ELSA analysis, a sample of 7181 adults (3325 males and 3856 females) was analyzed, with a mean age of 68.45 (SD = 10.13; age range = 50-99 years). This represents individuals who have taken part in the study between 2002/3 to 2018/19. Previous work has shown that the year/wave that the participant entered the study does not affect the relationship between polygenic prediction scores and frailty outcomes(4). Ethical approval for ELSA was gained via the South Central – Berkshire Research Ethics Committee (21/SC/0030, 22nd March 2021) and all methods were performed in accordance with the relevant guidelines and regulations. The ELSA genetics team approved a request for this study to utilize raw genotypes and provided linked genome-wide association study (GWAS) data to the phenotypic data curated at the Advanced Care Research Centre, University of Edinburgh(4).

***Prospective Imaging Study of Aging (PISA).*** PISA is a longitudinal cohort of ~3800 Australian adults aged 40-80 years(5). Since this cohort includes twin pairs, we selected one of each pair at random to be included in our current analyses. When the Frailty Index (FI) was derived and linked with genotypic data, 3265 participants (1034 males and 2231 females) remained for the main analysis, with a mean age of 60.34 (SD = 6.98). Participants are community dwelling and were recruited from a pool of participants who previously volunteered for research on risk factors for physical or psychiatric conditions and had genotypes already available. The PISA study protocol was approved by the Human Research Ethics Committee (HREC) at QIMR Berghofer Medical Research Institute.

#### Description of the Cohort Samples Used for Age-Stratified Sensitivity Polygenic Risk Score Analyses

To understand the effect of the polygenic risk scores (PRS) of the frailty latent factors as individuals age, we performed additional age-stratified analyses in each of the external cohorts. For LBC1936 we used data from waves 3 and 5 to utilize all the longitudinally measured waves in which the relevant data was present. The mean age at wave 3 was 76.30 (SD = 0.68; *N* = 649 (339 males, 310 females)) and was 82.06 at Wave 5 (SD = 0.53; *N* = 402 (196 males, 206 females)). We utilized these data for the univariate PRS analyses to predict the efficacy of the latent frailty factors in predicting Frailty Index (FI) and Fried Frailty Score (FFS) status in these waves of data, but we did not perform the age-stratified multivariate elastic net regression analyses with LBC1936 data owing to small sample sizes for waves 3 and 5.

To understand the effect of age on the prediction of FI status for individuals in ELSA and PISA, we split the data into 10-year age groups up to 90 years old in ELSA and up to 80 years old in PISA. In ELSA the 50-60 group mean age was 56.24 (SD = 2.81, *N* = 1788 (882 males, 906 females)). In PISA the 50-60 group mean age was 55.61 (SD = 3.03, *N* = 1455 (415 males, 1040 females)). In ELSA the 60-70 group mean age 65.59 (SD = 2.84, *N* = 2543 (1192 males, 1351 females)). In PISA the 60-70 group mean age was 64.99 (SD = 2.71, *N* = 1341 (431 males, 910 females)). In ELSA the 70-80 group mean age was 75.03 (SD = 2.85, *N* = 1912 (931 males, 981 females)). In PISA the 70-80 group mean age was 72.33 (SD = 1.41, *N* = 271 (102 males, 169 females)). In ELSA the 80-90 group mean age was 84.15 (SD = 2.55, *N* = 827 (341 males, 486 females)). We ran the univariate PRS regression analyses in each of these age groups, but we did not include the 70-80 years age group from PISA in the multivariate elastic net regression analyses because this sample was too under-powered.

We additionally ran both the univariate regression PRS analyses and the multivariate elastic net regression PRS analyses on a subset of the main ELSA and PISA samples that were aged ≥ 65 years old (PISA: mean age = 68.55, SD = 2.82, *N* = 991 (372 males, 619 females); ELSA: mean age = 68.55, SD = 2.82, *N* = 4407 (2066 males, 2341 females)).

**Description of the Frailty Outcomes Used for Polygenic Risk Score Prediction in ELSA, PISA and LBC1936**

***Frailty Index (FI).*** This measure was created in each of the external cohorts (LBC1936, ELSA and PISA) and includes frailty deficits that cover physical, biological, social, psychological and cognitive domains. The list of the included deficits in each FI are available in **Tables S52-54**. Deficits were either binary (coded as 0 (absent) or 1 (present) or 0.5 to represent a partially present deficit) or were continuous on a scale ranging from 0 to 1. The number of deficits was summed then divided by the total number of measured deficits to create a proportional FI score for each individual. Resulting FI scores ranged from 0 to 1, with higher scores indicating a higher frailty level. Our FI phenotypes were analyzed using linear regression.

***Fried Frailty Score (FFS).*** The FFS was only available in LBC1936 and was formed from five physical health traits: weight loss, exhaustion, physical inactivity, walking speed, and grip strength. If an individual did not display any of these 5 traits they were defined as healthy controls, whereas cases included individuals who were classed as being either pre-frail (i.e. the presence of 1-2 of these traits) or frail (i.e. if 3 or more traits were present) (6). We used the FFS from waves 1, 3 and 5 in our analyses to assess age-specific differences in physical frailty prediction. This resulted in 483 healthy controls, 514 pre-frail individuals and 94 frail individuals in the Wave 1 sample (used in the main PRS analyses), 268 healthy controls, 327 pre-frail individuals and 102 frail individuals in the Wave 3 sample, and 156 healthy controls, 214 pre-frail individuals and 61 frail individuals in the Wave 5 sample (used in the age-stratified sensitivity PRS analyses). We analyzed the FFS phenotype using multinomial logistic regression using non-frail individuals as the reference group.

**Description of the Frailty-Related Health Outcomes Used for Polygenic Risk Score Prediction**

***Cognitive Ability and Cognitive Change.*** Cognitive ability was measured in LBC1936. This measure was derived from a rich battery of 13 validated cognitive tests administered by psychologists at the initial wave of testing (Wave 1), where participants were ~70 years old. The cognitive tests included Matrix Reasoning, Block Design, Spatial Span, National Adult Reading Test, Wechsler Test of Adult Reading, Verbal Fluency, Verbal Paired Associates, Logical Memory, Digit Span Backwards, Symbol Search, Digit Symbol, Inspection Time and Choice Reaction Time. These were repeated at each subsequent wave (Wave 2: mean age = 72.55, SD = 0.68, *N* = 649 (339 males, 310 females); Wave 3: mean age = 76.30, SD = 0.68, *N* = 649 (339 males, 310 females); Wave 4: mean age = 79.39, SD = 0.62, *N* = 514 (216 males, 298 females) and Wave 5: mean age = 82.06, SD = 0.53, *N* = 402 (196 males, 206 females)). We used the results for all 13 tests to apply latent growth curve (LGC) modelling (i.e., a Factor of Curves model with full information maximum likelihood estimation) to estimate each individual's cognitive ability at Wave 1 (i.e. the intercept estimate; mean age = 69.60) and cognitive change across all waves (i.e. the slope estimate; age 70–82)(7). Individuals were retained in the analysis if they only had baseline data available since the estimates for intercept (cross-sectional) and slope (longitudinal) associations are derived simultaneously from the same model using all available data(8). We then used the *lavpredict* function in the lavaan R package to extract continuous intercept (cognitive ability) and slope (cognitive change) scores for each participant. We used linear regression to assess the association between these outcomes and the frailty PRS's.

***Motoric Cognitive Risk (MCR).*** MCR has been previously derived in LBC1936 (Wave 3) and is a measure that is based on subjective cognitive complaints and objective slow gait with no diagnosed disability or dementia present(9). We defined slow gait speed as walking speed that was one standard deviation below age- and sex-matched means. In LBC1936, this was calculated by digitally timing participants to walk 6 meters. If a participant confirmed that they had problems with memory, in response to the question "Do you currently have any problems with your memory?", this was recorded as a subjective cognitive complaint. Presence of disability or dementia was defined as present if an individual had a Townsend Disability score over 1.5 standard deviation from the mean, a confirmed dementia diagnosis and/or a Mini-Mental State Examination (MMSE) score of < 24/30. To investigate whether any of the frailty PRS's predicted MCR in LBC1936, we used competing risk regression models to examine the risk of developing MCR when death was a competing risk.

***Mortality.*** We used data from the General Register Office for Scotland to measure deaths in the participants in LBC1936, which we coded as a binary variable (0/1). We included both age in days at death and age at last censor to measure survival via both death and dropout. We conducted survival analysis using Cox proportional hazard regression models to examine the association between time to death and the frailty PRS's. We extracted hazard ratios and standardized beta coefficients from the model to compare effect sizes easily with other outcomes.

***Stroke.*** We derived the binary stroke measures for LBC1936, ELSA and PISA from self-report items where participants were asked whether they had ever had a stroke or a transient ischemic attack (TIA). We used logistic regression to assess the association between stroke and the frailty PRS outcomes in PISA and ELSA. We used a competing risk regression model to test for the association between stroke and frailty PRS with dropout as a competing risk in LBC1936 (Wave 1).

***Memory Problems.*** We utilized the self-report binary variable for memory complaints in PISA as an outcome for memory disturbances, which was derived by asking participants if they "suffer from memory complaints". We used logistic regression to measure the association between memory and the frailty PRS outcomes.

***Dementia.*** We created a binary variable for self-reported incident dementia in the ELSA dataset and analyzed the association between dementia and the frailty PRS outcomes using logistic regression.

### Supplementary Results

#### Linkage Disequilibrium Score Regression to Assess Genomic Signal of the Latent Frailty Factors

Quantile-Quantile plots of the SNP effects revealed clear genetic signal across all latent frailty factors. Results from linkage disequilibrium score regression (LDSC) further revealed that this reflected meaningful polygenic signal for each latent factor with minimal bias from population confounding, as evidenced by mean $\chi^2$ statistics and $\lambda_{GC}$ substantially greater than the LD intercept (**Supplementary Figure 1 and Table 1**).

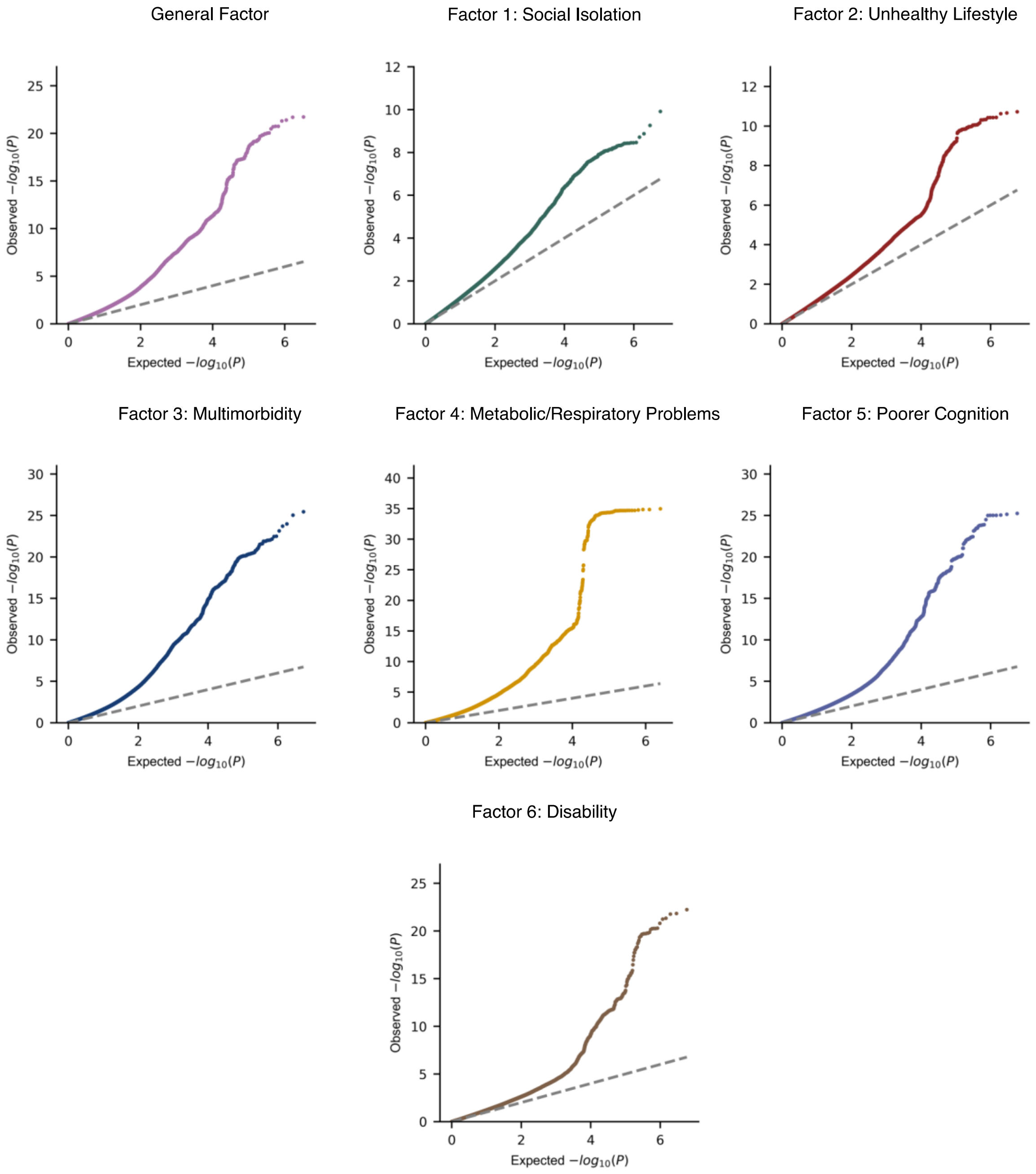


**Supplementary Figure 1: Quantile-Quantile (QQ) plots of the measured SNPs for each latent frailty factor GWAS.** For all latent frailty factors, there was substantial inflation of the quantile distribution of observed p-values versus the quantile distribution of the expected (i.e. null) p-values.

**Table 1: Results from linkage disequilibrium score regression of the GWAS results for each frailty latent factor conducted in Genomic SEM.** SE = standard error; LDSC = linkage disequilibrium score regression.

| Latent Frailty Factors | Mean $\chi^2$ | $\lambda$ Genomic Control | LDSC intercept (SE) |
|---|---|---|---|
| General Factor | 1.98 | 1.78 | 1.027 (0.017) |
| Factor 1 (social isolation) | 1.34 | 1.29 | 1.046 (0.010) |
| Factor 2 (unhealthy lifestyle) | 1.28 | 1.23 | 0.991 (0.009) |
| Factor 3 (multimorbidity) | 2.17 | 1.84 | 1.178 (0.017) |
| Factor 4 (metabolic/respiratory problems) | 2.40 | 2.05 | 1.125 (0.024) |
| Factor 5 (poorer cognition) | 1.81 | 1.63 | 1.102 (0.014) |
| Factor 6 (disability) | 1.36 | 1.30 | 1.069 (0.010) |

### Gene Prioritization Results for the Latent Frailty Factors

We used 5 gene mapping methods (positional, eQTL, chromatin interaction, MAGMA gene-based analysis and summary-data based Mendelian randomization) to identify genes that were potentially driving the shared genetic effects underlying each latent frailty factor. Any gene that was mapped to a latent factor by three or more of these methods was included in the pathway analysis outlined in the main manuscript. **Supplementary Figure 2** displays the number of genes mapped by different methods for the General Factor of frailty, and **Supplementary Figure 3** shows how many genes were mapped by the different methodologies for the latent residual factors of frailty (i.e., Factors 1 to 6).

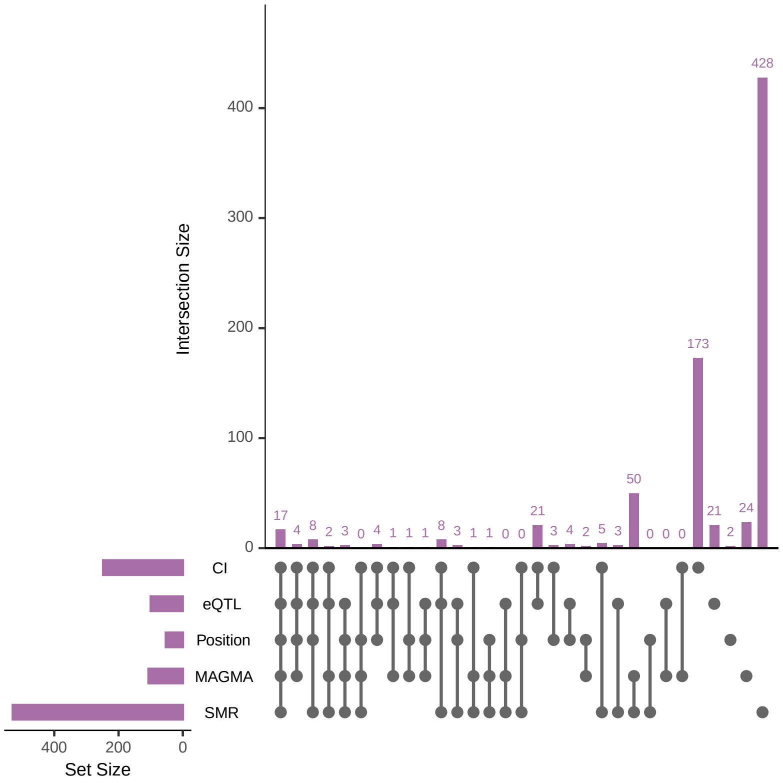


**Supplementary Figure 2: An UpSet plot displaying the number of genes mapped to the General Factor of frailty by the 5 different gene mapping methodologies.** CI = chromatin interaction; eQTL = expression quantitative trait locus; MAGMA = Multi-marker analysis of genomic annotation gene analysis; SMR = summary data-based Mendelian randomization.

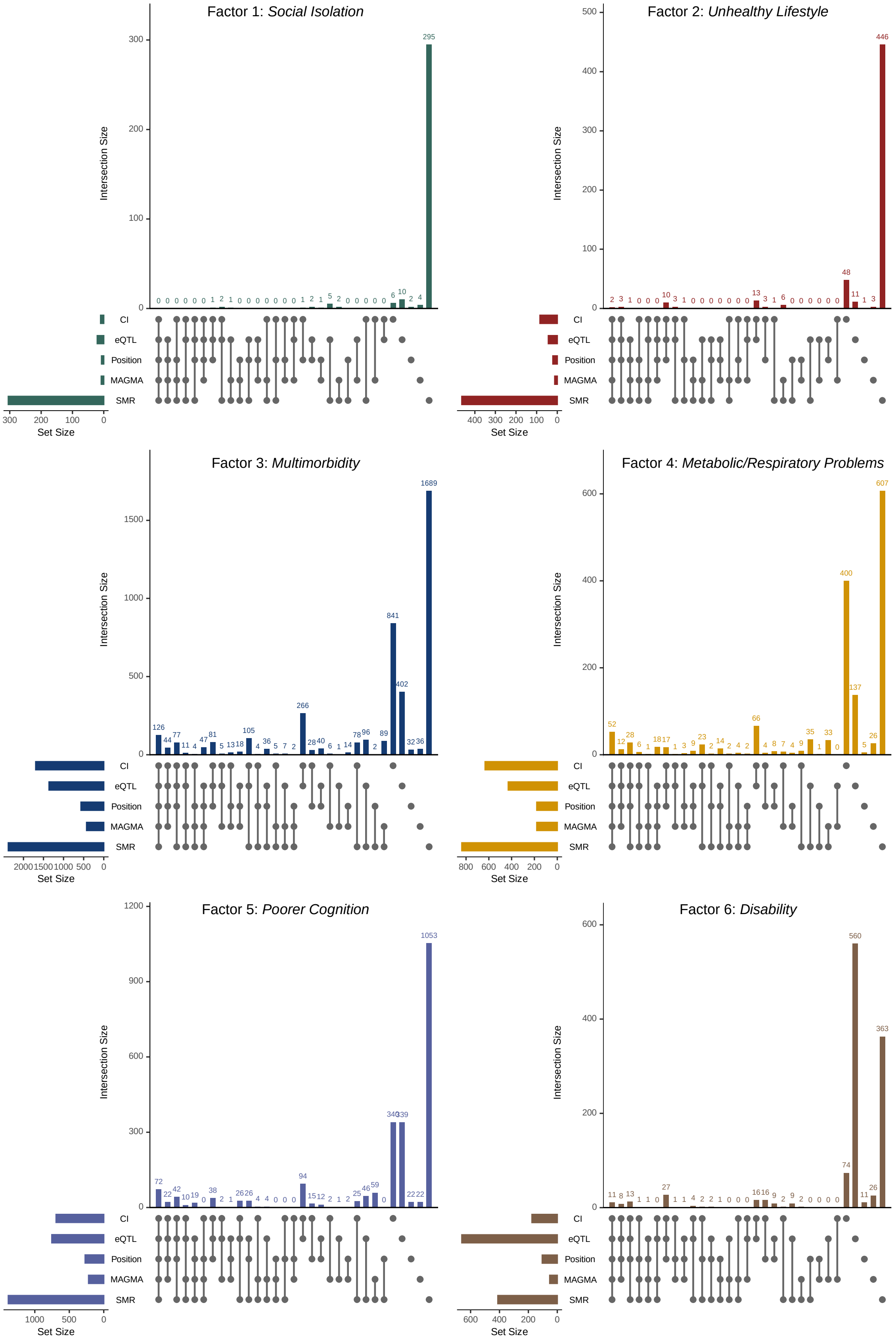


**Supplementary Figure 3: UpSet plots displaying the number of genes mapped to the latent residual factors of frailty (i.e., Factors 1-6) by the 5 different gene mapping methodologies.** CI = chromatin interaction; eQTL = expression quantitative trait locus; MAGMA = Multi-marker analysis of genomic annotation gene analysis; SMR = summary data-based Mendelian randomization.

#### Polygenic Risk Scores to Predict Fried Frailty Score Status in LBC 1936

As detailed in the main manuscript, we calculated polygenic risk scores (PRS) for each latent frailty factor and used multiple regression to combine these scores into a "Multi-PRS" that captured the genetic variance across all of the latent factors in our model. The Fried Frailty Score PRS (FFS-PRS) was the strongest predictor of the FFS for Wave 1 of LBC1936, which is to be expected given that it is a PRS constructed from the same phenotype (**Supplementary Figure 4** and **Table S44**). Despite this predictive advantage for the FFS-PRS, we found that the PRS for the General Factor of frailty and our Multi-PRS were the most predictive of the FFS outcome at Wave 3 of LBC1936 when we conducted our age-stratified sensitivity analyses in this cohort (**Supplementary Figure 4** and **Table S44**).

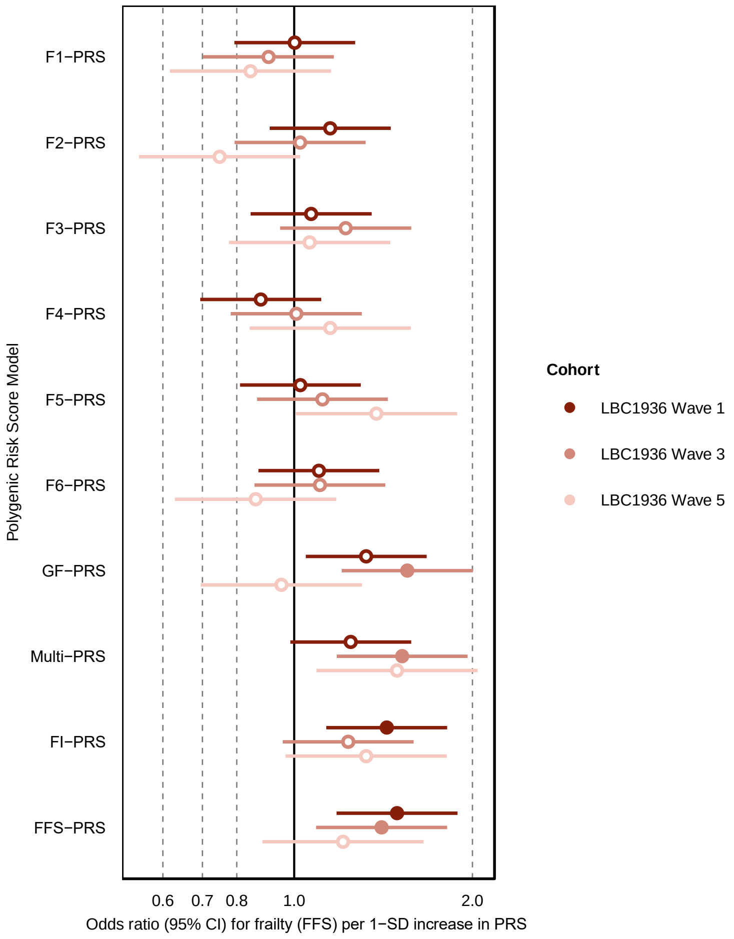


**Supplementary Figure 4: Results from the polygenic risk score prediction of the Fried Frailty Score (FFS) in the Lothian Birth Cohort 1936 (LBC1936) Waves 1, 3 and 5.** We display a forest plot comparing the odds ratios (95% Confidence Intervals [CI]) for each standard deviation (SD) increase in frailty (measured using the FFS) for each of our standardized frailty PRS phenotypes in each measured wave of data in LBC1936. Significant predictions (i.e. $P_{FDR} < 0.05$) are depicted as filled in dots, whereas non-significant predictions are depicted as an empty circle. CI = confidence intervals; SD = standard deviation. GF = General Factor; F1-6 = Factors 1-6.

#### Elastic Net Regression to Rank the Contributions of Each Latent Frailty Factor in the Prediction of Frailty Status in ELSA, PISA and LBC 1936

We conducted an elastic net regression analysis to rank the contributions of the PRS for each of the seven latent frailty factors in predicting frailty status (measured by either the FI or the FFS) in ELSA, PISA and LBC1936 Wave 1 (**Supplementary Figure 5**). We found evidence to support the use of all seven latent frailty factors, at differing levels of prediction, for assessing frailty status in external data, with the General Factor of frailty showing the most consistent effects across cohorts.

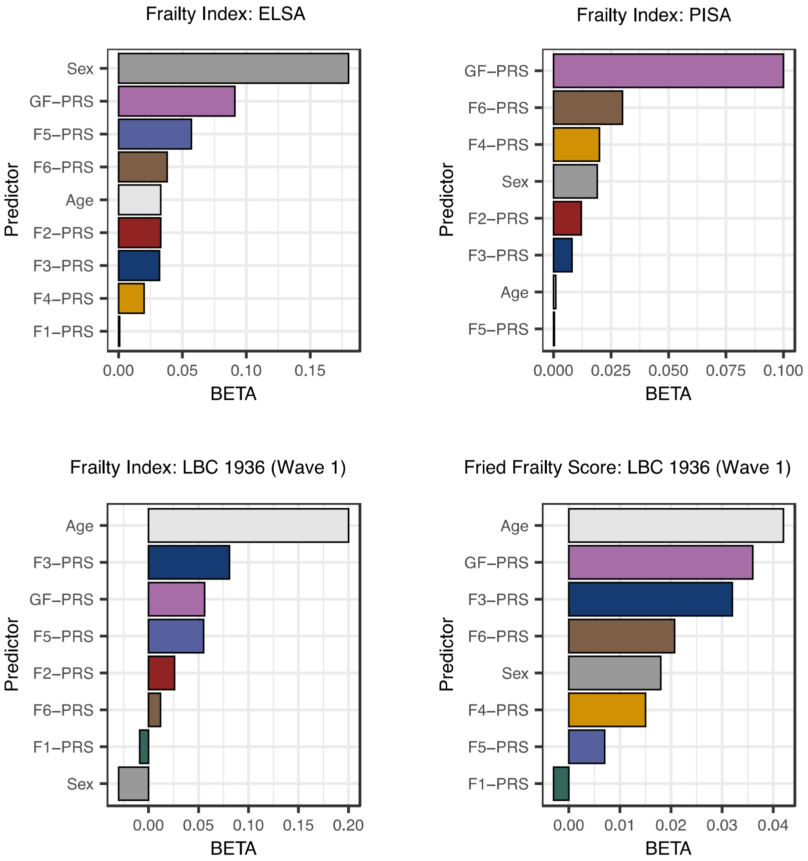


**Supplementary Figure 5: Standardized results of the elastic net regression analysis ranking the contributions of the seven latent frailty factors in predicting frailty status in external data.** Only predictors that contributed significant distinct variance in predicting the labelled outcome were retained in the final model results and are displayed in these plots. Each model included all seven latent factor PRS's as well as age, sex and ancestral principal components as covariates. PRS = polygenic risk score; GF = General Factor; F1-6 = Factors 1-6; LBC1936 = Lothian Birth Cohort 1986; PISA = Prospective Imaging Study of Aging; ELSA = English Longitudinal Study of Aging.

### Results of the Sensitivity Analyses that Examined Age-Stratified Polygenic Prediction of Frailty in ELSA, PISA and LBC 1936

To assess the prediction of the PRS's for each of our latent frailty factors at different ages, we reran the same multiple regression and elastic net regression analyses as we did in the main analysis in age-stratified samples for ELSA and PISA and data from waves 3 and 5 of LBC1936 (**Supplementary Methods**). We found that the PRS for the General Factor of frailty provided better prediction at younger ages across all cohorts (**Supplementary Figure 6** and **Supplementary Figure 7**). In contrast, the Multi-PRS (i.e. the PRS of the combined effect of all seven latent frailty factors) demonstrated more stable prediction across age groups, both in terms of the amount of variance it explained (**Supplementary Figure 6**) and because it was the only PRS to remain significantly predictive $(P_{FDR} < 0.05)$ of frailty status (measured using the FI) in all the measured age groups across all three cohorts (**Supplementary Figure 7**). Furthermore, as we demonstrated in the main analysis, the Multi-PRS of frailty outperformed the FI-PRS and FFS-PRS estimated from prior aggregate GWAS phenotypes, indicating its utility in estimating frailty status across different age ranges in adults aged over 50 years (**Supplementary Figure 7**).

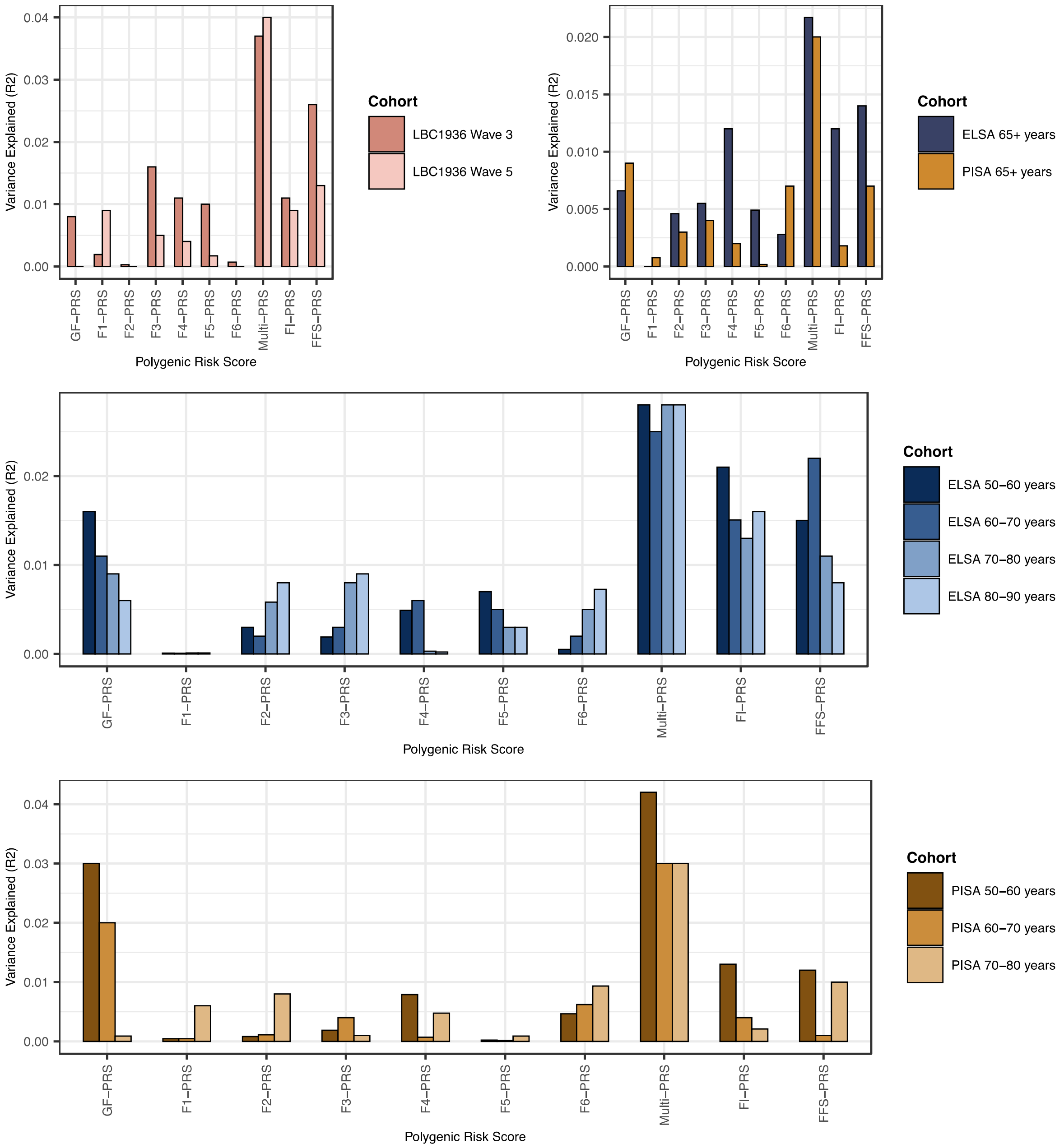


**Supplementary Figure 6: Variance explained (R2) by each frailty polygenic risk score (PRS) for predicting frailty status in external samples stratified by age.** Each grouped bar plot denotes the amount of variance explained (R2) by each of the standardized frailty PRS's that we created for predicting frailty status (measured by the Frailty Index) in all three external datasets. The samples were split by age group (ELSA and PISA) or by longitudinal wave (LBC1936) as noted in the key for each plot. LBC1936 = Lothian Birth Cohort 1936; PISA = Prospective Imaging Study of Aging; ELSA = English Longitudinal Study of Aging; F1-F6 = Factors 1-6; GF = General Factor; FI = Frailty Index; FFS = Fried Frailty Score.

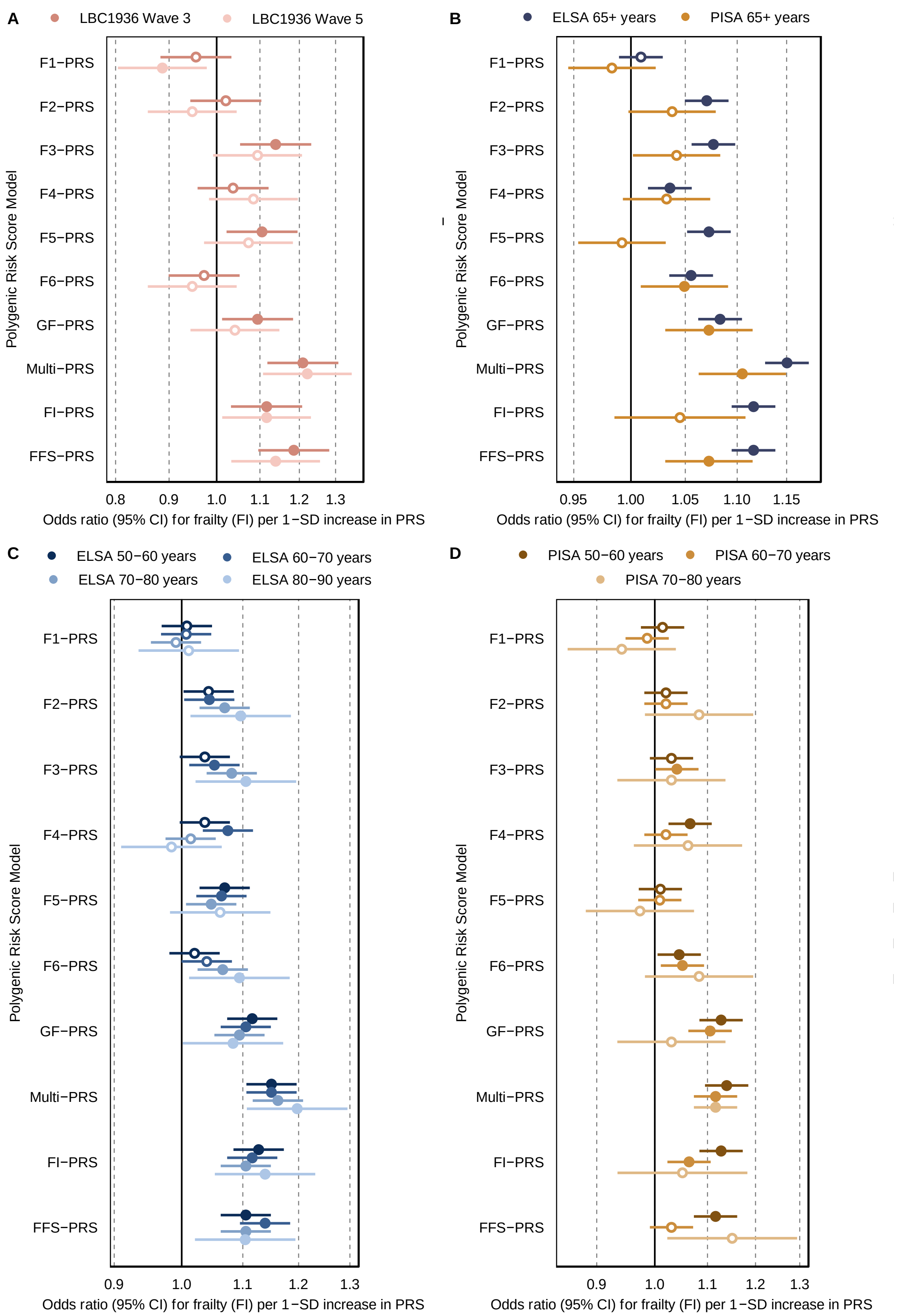


**Supplementary Figure 7: Forest plots of the association between Frailty Index status and our standardized frailty PRS phenotypes in the age-stratified samples from external cohorts.** Values are displayed as odds ratios (95% Confidence Intervals [CI]) for each standard deviation (SD) increase in frailty. Significant predictions ($P_{FDR} < 0.05$) are depicted as filled in dots. LBC1936 = Lothian Birth Cohort 1936; PISA = Prospective Imaging Study of Aging; ELSA = English Longitudinal Study of Aging; F1-F6 = Factors 1-6; GF = General Factor; FI = Frailty Index; FFS = Fried Frailty Score; PRS = polygenic risk score.

We then used elastic net regression to compare the predictive performance of our Multi-PRS against the aggregate FI-PRS and FFS-PRS measures in a model that also included age, sex and ancestral principal components as covariates, which we tested in each of the age groups in ELSA and PISA (except for the 70-80 years age group in PISA owing to small sample size) (**Supplementary Figure 8**). We found that the Multi-PRS provided greater prediction than either of the two aggregate PRS measures across all the tested age groups in ELSA and PISA. Furthermore, the accuracy of the Multi-PRS for predicting FI status in participants remained stable among age groups in both ELSA ($\beta$ range = 0.094-0.11) and PISA ($\beta$ range = 0.07-0.11), whereas age became more predictive with increasing age (ELSA $\beta$ range = 0.009-0.061; PISA $\beta$ range = 0-0.035), which is to be expected because age is strongly associated with frailty risk. We also identified a large predictive effect of sex on frailty status in ELSA that increased by age group ($\beta$ range = 0.076-0.30), but not in PISA ($\beta$ range = 0.012-0.015), although this difference in effect may be owing to differences in the sex ratio between these two samples (PISA has ~50% more females than males, whereas ELSA is approximately equal (**Supplementary Methods**)).

Finally, we used elastic net regression in the ELSA and PISA samples grouped by age to rank the predictive contributions of the PRS for each of the seven latent frailty factors from our model, when sex, age and ancestral principal components were also included as covariates. The PRS for the General Factor of frailty was the best predictor of FI status across all of the age groups in ELSA and PISA, except for the 80–90-year-old age group in ELSA where the PRS for Factor 3 (multimorbidity) provided greater prediction (**Supplementary Figure 9**). Similarly to our main results, we also found that there was evidence of significant unique predictive contributions of the PRS's for all six latent residual factors of frailty (i.e., Factors 1-6), but their ranking and predictive contribution varied across age groups (**Supplementary Figure 9**). This suggests that these subsets of frailty deficits may exert more time-specific influences on frailty onset compared to the core frailty pathways underpinning the General Factor. This could have implications for the development of future treatments or prevention strategies for frailty because certain deficit pathways may require intervention at more precise timepoints during the life course.

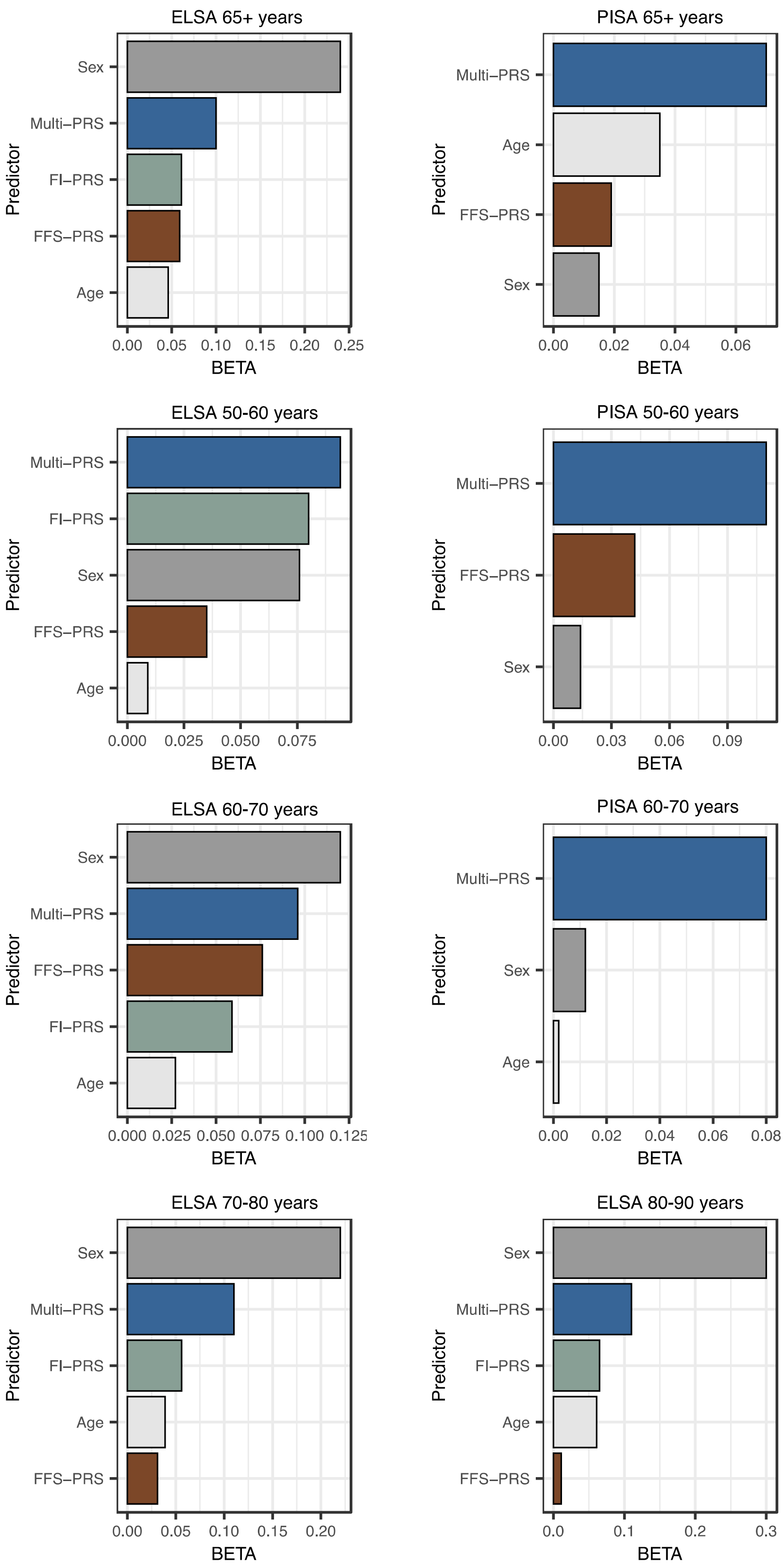


**Supplementary Figure 8: Standardized results from the age-stratified elastic net regression analyses for Frailty Index prediction in the PISA and ELSA cohorts.** These analyses ranked the performance of our Multi-PRS (i.e. combined latent frailty factor score) when modelled with the aggregate Frailty Index PRS (FI-PRS), the aggregate Fried Frailty Score PRS (FFS-PRS), age, sex and ancestral principal components as covariates. Only the predictors that explained significant additional unique variance over and above the other covariates are included in these plots. PISA = Prospective Imaging Study of Aging; ELSA = English Longitudinal Study of Aging; PRS = polygenic risk score.

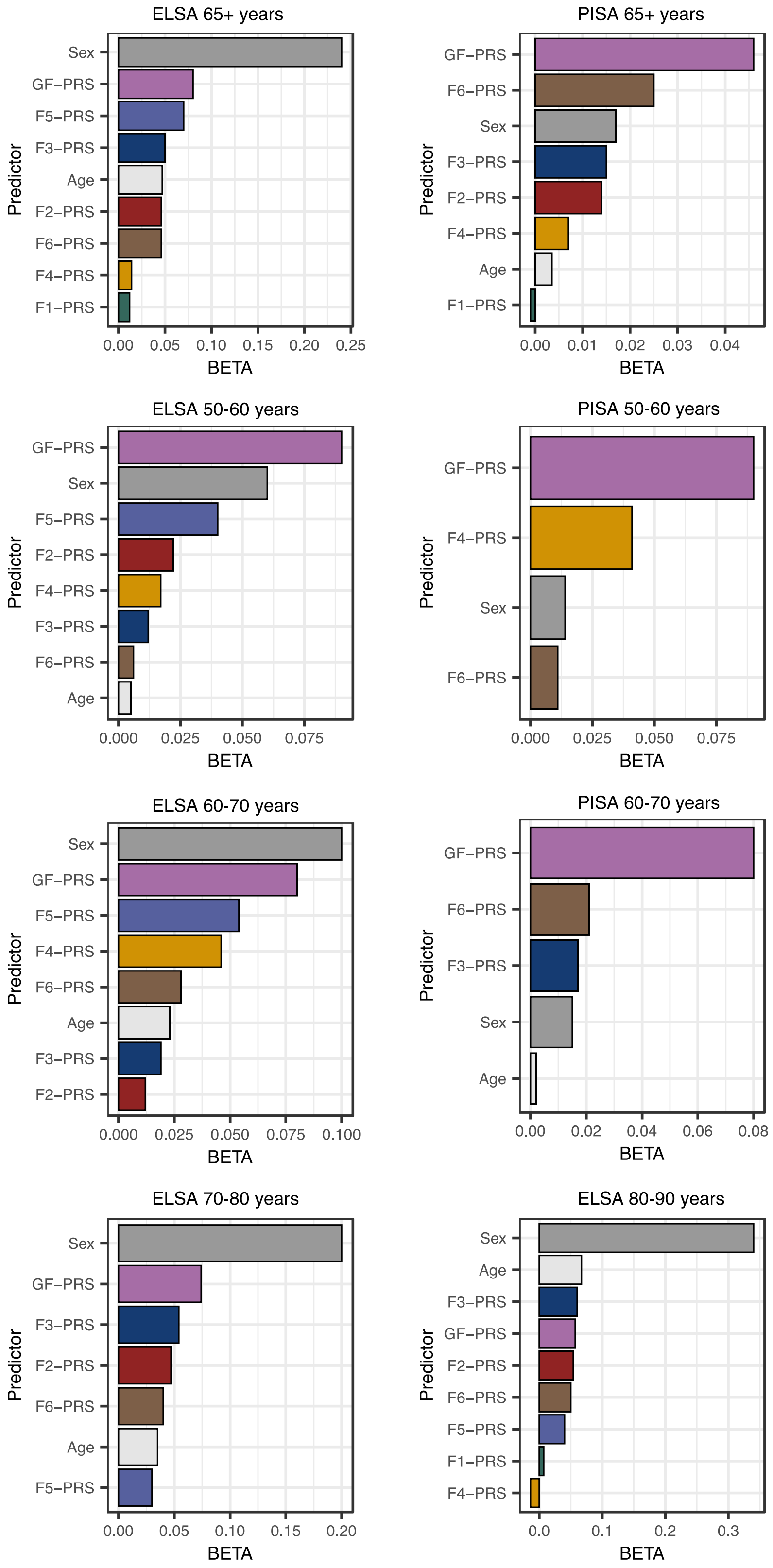


**Supplementary Figure 9: Standardized results of the age-stratified elastic net regression analysis ranking the contributions of the seven latent frailty factors in predicting Frailty Index status in ELSA and PISA.** Only predictors that contributed significant distinct variance in predicting Frailty Index status were retained in the final model results and are displayed in these plots. Each model included all seven latent factor PRS's as well as age, sex and ancestral principal components as covariates. PRS = polygenic risk score; GF = General Factor; F1-6 = Factors 1-6; PISA = Prospective Imaging Study of Aging; ELSA = English Longitudinal Study of Aging.

### Simulation Results of New Method to Calculate $Q_{SNP}$ Heterogeneity Index

As we outlined in the main text, we developed a more computationally tractable method to calculate the factor-specific $Q_{SNP}$ heterogeneity index for each measured SNP in a multivariate GWAS conducted within GenomicSEM. To validate this new method, we simulated 3 different factor models, including a 2-factor model with three indicators on each factor (2 degrees of freedom [df]), a 2-factor model with four indicators on each factor (3 df) and a 2-factor model with six indicators on each factor (5 df). **Table 2** and **Supplementary Figure 10** provide evidence that there was not a significant difference in the mean $Q_{SNP}$ estimates between the new versus the old method and that our new method remained $\chi^2$-distributed at a comparable level to the old method. **Table 3** further demonstrates that the new method produced a slightly lower type 1 error rate compared to the old method.

**Table 2: Comparisons of the new and old $Q_{SNP}$ estimation methods from our simulation tests.** SD = standard deviation; df = degrees of freedom.

| **Simulation Model** | **New Method** | | **Old Method** | | **Mean difference between new and old method** |
|---|---|---|---|---|---|
| | *Mean $Q_{SNP}$ estimate* | *SD of $Q_{SNP}$ estimate* | *Mean $Q_{SNP}$ estimate* | *SD of $Q_{SNP}$ estimate* | |
| 2 df model | 2.004 | 1.939 | 2.049 | 1.984 | -0.044 |
| 3 df model | 3.050 | 2.503 | 3.099 | 2.543 | -0.049 |
| 5 df model | 4.951 | 3.099 | 5.009 | 3.146 | -0.058 |

**Table 3: Comparison of type-1 error rates between the new and old $Q_{SNP}$ estimation methods.** Df = degrees of freedom.

| **Simulation Model** | **New Method** | | | **Old Method** | | |
|---|---|---|---|---|---|---|
| | *Number of type 1 errors* | *Number of non-type-1 errors* | *Ratio* | *Number of type 1 errors* | *Number of non-type-1 errors* | *Ratio* |
| 2 df model | 46 | 954 | 0.046 | 49 | 951 | 0.049 |
| 3 df model | 49 | 951 | 0.049 | 52 | 948 | 0.052 |
| 5 df model | 41 | 959 | 0.041 | 47 | 953 | 0.047 |

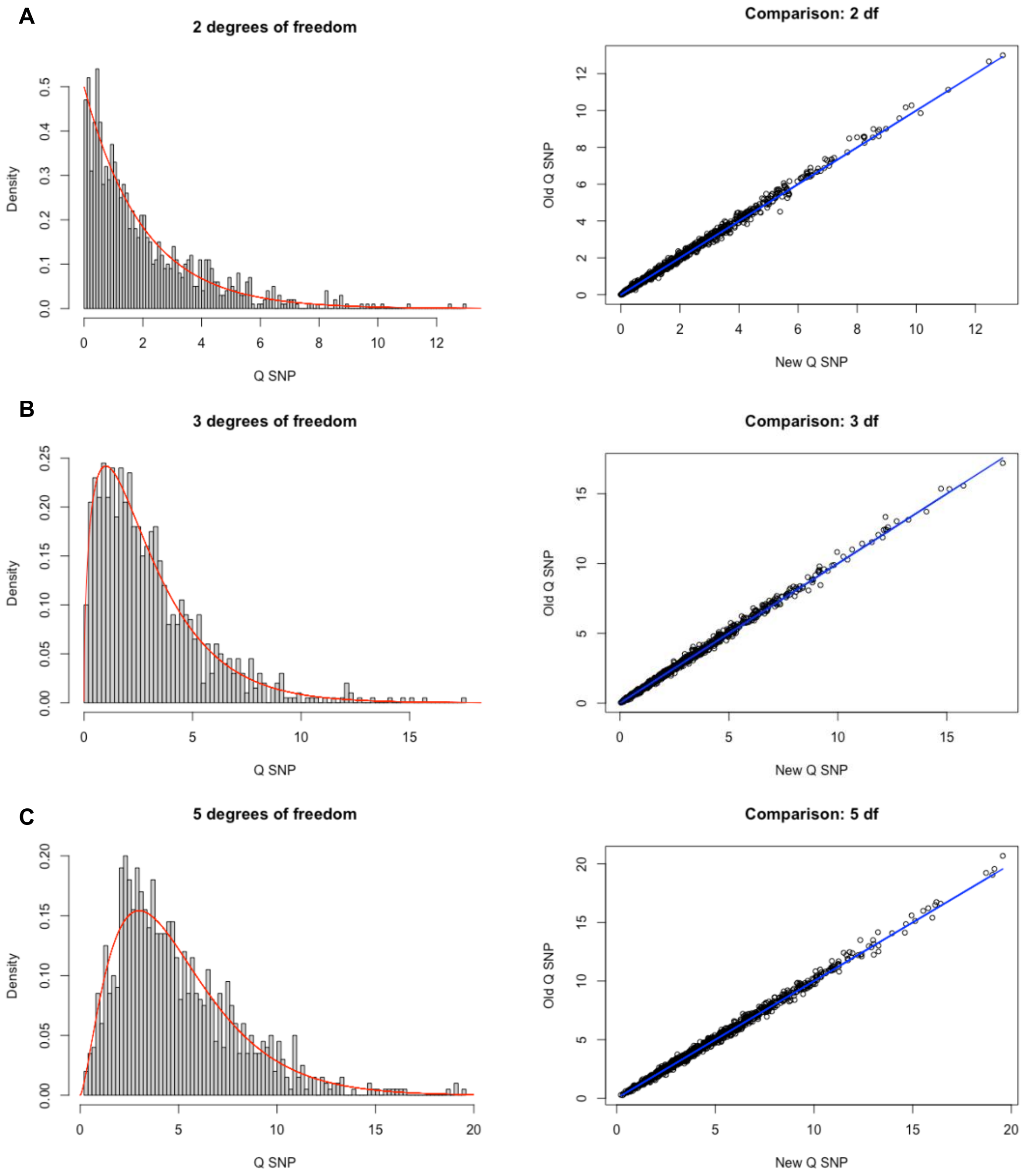


**Supplementary Figure 10: Distributions of the $Q_{SNP}$ $\chi^2$ statistics of our simulated models.** Left-hand plots depict histograms of the distributions for the simulated $Q_{SNP}$ $\chi^2$ statistics generated using the new estimation method within the GenomicSEM R package and the right-hand plots present scatter plots of the $Q_{SNP}$ $\chi^2$ statistics generated from the old estimation method on the y axis and the new estimation method on the x axis. *Panel A* are results for the 2df model (2 factors with 3 indicators), *Panel B* shows the 3df model results (2 factors with 4 indicators) and *Panel C* shows the 5df model results (2 factors with 6 indicators).
